# Nocturnal and Diurnal Measures of Autonomic Function in Idiopathic Hypersomnia and Type 1 Narcolepsy

**DOI:** 10.64898/2026.04.09.26349889

**Authors:** Jennifer Zitser, Luca Baldelli, Hash Brown Taha, Oliver Sibal, Giacomo Chiaro, Annagrazia Cecere, Giorgio Barletta, Pietro Cortelli, Pietro Guaraldi, Mitchell G. Miglis

**Affiliations:** Department of Psychiatry and Behavioral Sciences, Division of Sleep Medicine, Stanford University, Palo Alto, CA, USA; Sleep Department, Neurology Department, Tel Aviv Sourasky medical center, Israel; IRCCS Istituto delle Scienze Neurologiche di Bologna, UOC Clinica Neurologica NeuroMet, Ospedale Bellaria, Via Altura 3, 40139, Bologna, Italy; Department of Biomedical and Neuromotor Sciences (DIBINEM), University of Bologna, Bologna, Italy; Washington University School of Medicine, St. Louis, MO, USA; Department of Neurology and Neurological Sciences, Stanford University, Palo Alto, CA, USA; National Autonomic Centre, The National Hospital for Neurology and Neurosurgery, London, United Kingdom

## Abstract

**Study Objectives:** Idiopathic hypersomnia (IH) is a central nervous system hypersomnia frequently accompanied by autonomic symptoms, yet objective physiological data are limited. We sought to characterize autonomic nervous system (ANS) dysfunction in IH using nocturnal heart rate variability (HRV) and diurnal autonomic reflex testing (ART), compared to individuals with type 1 narcolepsy (NT1) and healthy controls (HCs).

**Methods:** Twenty-four adults with IH, 10 with NT1, and 14 HCs underwent overnight video polysomnography with HRV analyses in time and frequency domains during stable slow-wave sleep and REM sleep. Comprehensive ART included sympathetic adrenergic (head-up tilt (HUT), Valsalva BP responses), parasympathetic cardiovagal (HRV to deep breathing, Valsalva ratio), and sudomotor (Q-Sweat) measures.

**Results:** IH participants were predominantly female, with over half reporting long sleep duration. Compared to NT1 and HC, participants with IH demonstrated a greater magnitude of orthostatic tachycardia on tilt (ΔHR 41.0 ± 16.3 vs. 26.3 ± 9.3 vs. 30.8 ± 9.3 bpm, p = 0.0086), as well as frequent sudomotor dysfunction (64.3%). IH participants demonstrated greater nocturnal and REM HR with reduced parasympathetic indices during REM, indicating diminished vagal modulation compared with HCs

**Conclusions:** IH is characterized by a distinct pattern of autonomic dysfunction, including pronounced orthostatic tachycardia, frequent sudomotor abnormalities, and reduced parasympathetic activity during sleep. These findings provide objective physiological evidence of ANS involvement in IH and delineate features that distinguish IH from NT1 and HCs.

## INTRODUCTION

Idiopathic hypersomnia (IH) is a central nervous system hypersomnia of unknown etiology characterized by excessive daytime sleepiness, unrefreshing sleep that tends to be long-lasting, sleep inertia and the absence of cataplexy.^1^ In addition to these cardinal symptoms, several studies have reported that patients with IH may exhibit signs and symptoms of autonomic nervous system (ANS) dysfunction. In Bedrich Roths’ seminal publication on IH that helped characterize the disorder, resting tachycardia was described in a significant number of patients.^2^ Subsequent publications have described other symptoms of ANS dysfunction including orthostatic intolerance, vasomotor instability, gastrointestinal upset, and temperature intolerance, among others.^3,4^ However, studies of objective autonomic dysfunction are limited, with one heart rate variability (HRV) study demonstrating increased parasympathetic activity during both wake and sleep in individuals with IH.^5^ While several studies have attempted to characterize objective measures of ANS dysfunction in narcolepsy,^6,7^ there are no studies, to our knowledge, that have attempted to accomplish this in IH. We previously reported a high burden of autonomic symptoms in IH, with worse symptoms directly correlating with worse daytime sleepiness, fatigue, and quality of life.^4^ We are following up on these findings in this study to better characterize ANS dysfunction in IH using objective nocturnal HRV measures as well as diurnal autonomic reflex testing (ART) in a cohort of participants with IH, compared to healthy controls (HC) and those with type-1 narcolepsy (NT1).

## METHODS

Adult participants ≥ 18 years of age with IH were consecutively recruited from the Stanford Sleep Disorders Center. Comparison groups (NT1 and HCs) were derived from a previously recruited cohort at the University of Bologna that also underwent detailed autonomic and sleeps assessments. Participants in the NT1 group and a subset of the HCs were included in a prior publication.^12^ The HC group consisted of healthy volunteers recruited at the University of Bologna for that study (n=7), as well as additional healthy volunteers (n=7) recruited from partners and acquaintances of patients attending the neurology clinic, using the same inclusion/exclusion criteria and acquisition procedures. None of the HCs had a sleep disorder or relevant psychiatric or medical condition. NT1 participants were recruited and evaluated at the University of Bologna.

Participants with IH and NT1 were diagnosed by the study investigators using current AASM criteria (mean sleep latency ≤ 8 minutes on MSLT with <2 sleep-onset REM periods (SOREMs) for IH and ≥ SOREMs for NT1).^13^ Participants were excluded if they had any pre-existing hypersomnia or autonomic disorder, or any untreated sleep disorder that, in the investigator’s opinion, would predispose participants to hypersomnia or autonomic dysfunction. All subjects gave written informed consent, and the study was approved by the local Institutional Review Boards of Stanford University and the University of Bologna.

In the Bologna cohort, NT1 participants were drug-free at the time of polysomnography (3/10 drug-naïve; 7/10 discontinued modafinil ≥2 weeks prior; 1/10 additionally discontinued clomipramine 1 month prior). HCs were drug-free except for two individuals (one taking ramipril, held during protocol days; one who discontinued prazepam 1 week prior and held paroxetine during protocol days). In the Stanford IH cohort, medications were also held for at least 5 half-lives prior to testing, consistent with the Bologna protocol. Prior to washout, wake promoting medications were used by 14/24 (58.3%) IH participants, 7/10 (70%) NT1 participants, and 1/14 (7.1%) HCs; autonomic medications were used by 9/24 (37.5%) IH participants, 0/10 NT1 participants, and 1/14 (7.1%) HCs; both sleep and autonomic medications were used by 7/24 (29.2%) IH participants and by no NT1 participants or HCs.

All subjects underwent overnight video polysomnogram (vPSG) at the Stanford Sleep Disorders Clinic or the University of Bologna. vPSGs at Stanford were acquired with the SOMNOmedics system according to AASM criteria.^13^ Apneas were defined as a ≥90 % reduction of airflow lasting ≥10 s. Hypopneas were defined as a ≥30 % reduction in nasal pressure signal excursions and associated ≥3 % desaturation or arousal. Sleep staging, arousals, and periodic limb movements were scored using the current AASM criteria.^13^ Data were collected and analyzed using DOMINO software (SOMNOmedics Inc.) vPSGs at the University of Bologna were acquired with Neurofax Electroencephalograph EEG-1200, Nihon Kohden Corp., Tokyo, Japan) and Albert Grass Heritage® Colleague TM PSG Model PSG16P-1, (Astro-Med, Inc, West Warwick, RI) and analyzed with Polysmith™ Sleep 9.0 (Nihon Kohden Corporation, Tokyo, Japan). The vPSG included continuous recording throughout one night of the following parameters: electroencephalogram (EEG: F3–A2, C3–A2, CZ–A1, O2–A1), right and left electrooculogram, electrocardiogram, electromyogram of the mylohyoideus, left and right anterior tibialis muscles, thoraco-abdominal breathing (strain gauge), and video-synchronized recording. All vPSGs were performed in acoustically isolated rooms, at constant temperature (22-24°C) and under the supervision of trained technicians. All vPSG channels were sampled at a frequency of 512Hz.

HRV analyses were performed on 5-min periods during stable slow wave sleep (SWS) and rapid eye movement (REM) sleep. After a stage change, only consecutive 5-min segments consisting of a particular stage were used for analysis. Preprocessing of the RR interval time series and computation of time and frequency domain measures of HRV were performed using Kubios HRV premium 3.0 software (Kubios Ltd., Kuopio, Finland) in accordance with standard guidelines^14^. The same investigator (J.Z.) visually selected the segments for HRV analysis. To be included in the analysis, segments had to be free from arousals, artifacts, sleep-disordered breathing events, body movements, and ectopic beats. Five-minute periods were selected for analysis in which the 30 s epoch before and after were artifact free to ensure a stationary signal.^14^

The square root of the mean of the squares of the successive differences between adjacent R-R intervals (RMSSD) was chosen as the time domain index of HRV, since it is strongly correlated with the parasympathetic modulation of HR.^14^ Spectral analysis of HRV was performed using a Fast Fourier Transform and autoregressive (AR) methods. Spectral power was calculated in the high frequency band (HF: 0.15–0.40 Hz) reflecting mostly parasympathetic activity, and in the low frequency band (LF: 0.04– 0.14 Hz) reflecting a combination of vagal and sympathetic activities.^14^ The LF/HF ratio was also calculated, but was interpreted cautiously and not as a simple index of sympathovagal balance.^14^ For the AR-based analyses, we derived absolute power in the very-low frequency, low-frequency and high-frequency bands (Abs. VLF AR, Abs. LF AR, Abs. HF AR) and their corresponding relative powers expressed as a percentage of total power (Rel. VLF AR, Rel. LF AR, Rel. HF AR).

All participants completed a battery of standardized diurnal ART, including sudomotor measures (Quantitative Sweat Measurement System (Q-sweat), WR Medical Electronics Co., Stillwater, MN), parasympathetic cardiovagal measures (HRV with deep breathing (HRDB), Valsalva HR ratio) and sympathetic adrenergic measures (Valsalva BP response, 10-minute head-up tilt (HUT) at an angle of 70 degrees). Beat-to-beat blood pressure (BP) was measured with finger plethysmography using a CNAP^®^ device (CN Systems Medizintechnik GmbH, Graz, Austria) and confirmed with an automated cuff sphygmomanometer over the brachial artery during HUT. Delta (Δ) HR was calculated as the difference in maximum and supine baseline HR measurements during HUT. Δ systolic BP (SBP) and diastolic blood pressure (DBP) was calculated as the difference between supine baseline and minimum SBP measurements during HUT.

Data was collected and analyzed using Testworks software (WR Medical Electronics). The overall integrated instrumental approach and laboratory procedures for autonomic nervous system assessment have been previously described.^15^

### Statistical analysis

For multi-group comparisons, one-way ANOVA or the Kruskal-Wallis test was used as appropriate, followed by post hoc analyses using Tukey’s HSD (equal variances), Games–Howell (unequal variances), or Dunn’s test for nonparametric data; the specific tests applied are indicated in the corresponding tables. For comparisons involving two groups, independent t-test or the Mann-Whitney U test, depending on data normality were conducted. Chi-square tests were used for categorical variables Receiver operating characteristic (ROC) analyses were conducted using binomial logistic regression models, with sensitivity and specificity calculated at the optimal cutoff maximizing Youden’s index. All statistical tests used for each analysis are indicated in the corresponding tables. All analyses and graphical visualizations were conducted in RStudio (version 2022.12.0+353).

## RESULTS

The multi-center cohort consisted of 24 participants with IH, 10 with NT1, and 14 HCs (Figure 1, Table 1). Participants with IH were predominantly female (22/24; 91.7%) compared to NT1 (1/10; 10%) and HCs (6/14; 57.1%, p < 0.0001), and had a lower body mass index (BMI) compared to NT1 participants. More than half of the IH cohort (14/24; 58.3%) reported long sleep (TST >11 h). As expected from the younger age of disease onset in the NT1 group, disease duration at the time of autonomic testing was substantially greater in NT1 compared to IH. All patients with NT1 were positive for HLA DQB1*06:02 and most (9/10; 90%) had low cerebrospinal fluid hypocretin-1 levels (not detectable in five participants; 17–92 pg/ml in four participants, 6,110 pg/ml in one participant).

**Table 1.** Demographic and clinical characteristics of patients with idiopathic hypersomnia (IH), narcolepsy (NC), and healthy controls (HC).

| Demographics | IH (n = 24)<br>Mean ± SD<br>Median<br>(IQR) | Narcolepsy (n = 10)<br>Mean ± SD<br>Median (IQR) | Controls (n = 14)<br>Mean ± SD<br>Median (IQR) | P-value | Post-hoc |
| --- | --- | --- | --- | --- | --- |
| Age (years) | 31.9 ± 7.7<br>31.5 (8.5) | 37.5 ± 7.6<br>35.5 (8.5) | 38.4 ± 10.3<br>36.5 (16.0) | 0.12 | — |
| Female/male (%) | 22/2 (91.7%) | 1/9 (10%) | 6/8 (57.1%) | <0.0001* | — |
| BMI (kg/m<sup>2</sup>) | 24.0 ± 4.6<br>23.8 (4.9) | 28.1 ± 3.6<br>27.8 (3.5) | 23.4 ± 4.4<br>23.4 (5.5) | 0.010* | NC vs. IH/HC |
| Disease onset age | 26.3 ± 10<br>26.5 (11) | 13.6 ± 5.6<br>13.5 (6.8) | — | — | — |
| Disease duration<br>(at autonomic testing) | 6.3 ± 7.7<br>2 (4) | 23.9 ± 10.5<br>21.0 (14.5) | — | — | — |
| Long sleep, n (%) | 14 (58.3%) | — | — | — | — |

Statistical tests were conducted using the were done using a one-way analysis of variance (ANOVA) or the Kruskal-Wallis test based on data distribution normality or chi-square test for categorical variables. Statistical significance (\*) is shown for P-value < 0.05. Long sleep was determined based on subjective reported total sleep time > 11 hours. Abbreviations: BMI – body mass index; SD – standard deviation; IQR – interquartile range.

**Figure 1.**
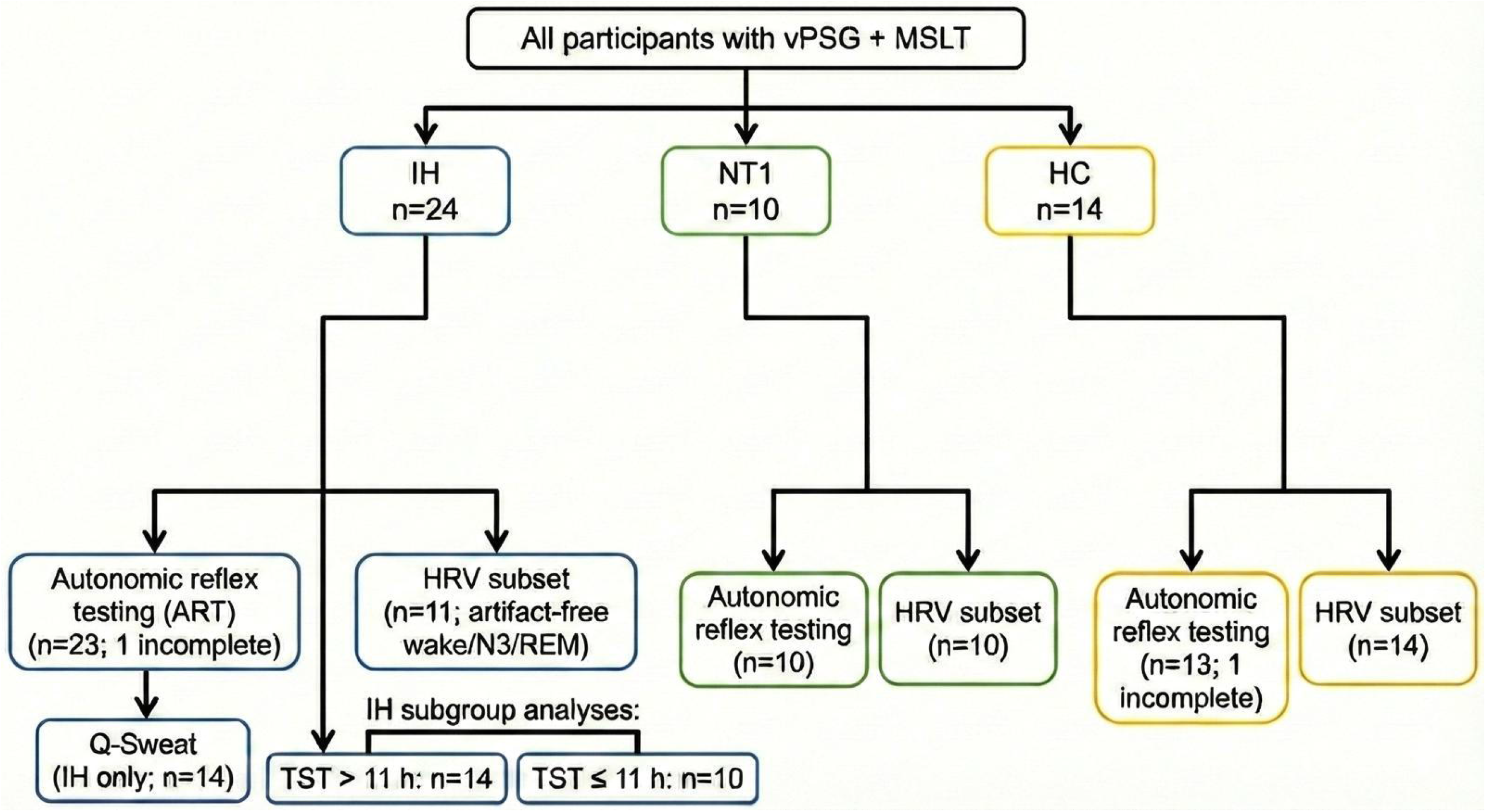
Flow diagram of study cohorts and analytic samples. The final cohort comprised 24 idiopathic hypersomnia (IH), 10 type 1 narcolepsy (NT1) and 14 healthy control (HC) participants, all of whom underwent PSG and MSLT. Boxes show how many from each group completed autonomic reflex testing (ART), sudomotor testing (Q-Sweat, IH only), nocturnal HRV analyses, and the IH subgroups with total sleep time > 11 h versus ≤ 11 h. Abbreviations: IH, idiopathic hypersomnia; NT1, type 1 narcolepsy; HC, healthy controls; vPSG, video polysomnography; MSLT, multiple sleep latency test; ART, autonomic reflex testing; HUT, head-up tilt; HRV, heart rate variability; TST, total sleep time.

### Diurnal Autonomic Reflex Testing

ART data were available for 23 IH participants and 13 HCs, as one IH participant and HC did not complete the full protocol (Table 2, Figure 2). For HRV analyses during PSG, artifact⍰free 5⍰minute segments of wake, N3, and REM were available in 11 IH, 10 NT1, and 14 HCs.

**Table 2.** Summary of autonomic function testing results in patients with idiopathic hypersomnia (IH), narcolepsy (NC), and healthy controls (HC).

| Diurnal Autonomic Variables | IH<br>(n = 23) | Narcolepsy<br>(n = 10) | Controls<br>(n = 13) | P value | Post-hoc |
| --- | --- | --- | --- | --- | --- |
| **Sympathetic adrenergic BP and HR on HUT and Valsalva Maneuver** |  |  |  |  |  |
| HUT SBP (mmHg): |  |  |  |  |  |
| Baseline | 123.3 ± 14.0 | 122.4 ± 15.8 | 123.1 ± 7.7 | 0.81<sup>b</sup> | — |
| 3 Min | 122.4 ± 16.0 | 131.3 ± 21.0 | 126.4 ± 9.8 | 0.33 |  |
| 10 Min<sup>1</sup> | 120.4 ± 17.3 | 128.0 ± 20.4 | 122.9 ± 7.8 | 0.57<sup>b</sup> |  |
| HUT DBP (mmHg)<sup>2</sup>: |  |  |  |  |  |
| Baseline | 74.8 ± 11.1 | 64.7 ± 8.7 | 67. ± 4.8 | 0.0077*<sup>b</sup> | IH vs. NC<sup>a</sup> |
| 3 Min | 79.4 ± 10.7 | 73.7 ± 12.0 | 75.7 ± 4.2 | 0.30<sup>b</sup> |  |
| 10 Min | 78.3 ± 11.9 | 73.4 ± 10.3 | 74.1 ± 5.4 | 0.37 |  |
| HUT HR (bpm): mean ± SD |  |  |  |  |  |
| Baseline | 78.4 ± 13.9 | 70.8 ± 9.3 | 67.6 ± 10.8 | 0.036* | IH vs. HC |
| 3 Min | 102.3 ± 18.1 | 83.5 ± 12.9 | 84.9 ± 13.3 | 0.0017* | IH vs. NC/HC |
| 10 Min<sup>1</sup> | 104.6 ± 18.4 | 84.7 ± 9.2 | 84.2 ± 12.8 | <0.001* | IH vs. NC/HC |
| HUT ΔSBP 3 minutes (mmHg) | -0.83 ± 10.8 | 8.9 ± 10.5 | 3.3 ± 7.8 | 0.044* | IH vs. NC |
| HUT ΔDBP 3 minutes (mmHg)<sup>2</sup> | 4.6 ± 7.6 | 9.0 ± 6.2 | 8.1 ± 3.7 | 0.12<sup>b</sup> | — |
| HUT ΔHR<sup>1</sup> | 41.0 ± 16.3 | 26.3 ± 9.3 | 30.8 ± 9.3 | 0.0086*<sup>b</sup> | IH vs. NC |
| Abnormal HUT: n (%) | 16 (69.6%) | 10 (100%) | 0 (0%) | — | — |
| Valsalva baseline to P2 minimum (mmHg)<sup>3</sup> | -12.9 ± 38.2 | — | — | — | — |
| Valsalva P2 recovery (mmHg)<sup>3</sup> | 11.4 ± 30.4 | — | — | — | — |
| Valsalva P4 overshoot (mmHg)<sup>3</sup> | 31.2 ± 20.2 | 41.4 ± 20.2 | 40.3 ± 18.4 | 0.35<sup>b</sup> | — |
| **Cardiovagal HR variables** |  |  |  |  |  |
| HRVdb (bpm)<sup>1</sup> | 23.3 ± 8.4 | 24.1 ± 7.8 | 26.8 ± 8.3 | 0.46 | — |
| Valsalva Ratio<sup>4</sup> | 2.2 ± 0.50 | 1.7 ± 0.36 | 1.9 ± 0.23 | 0.011<sup>*a</sup> | IH vs. NC |
| Abnormal HRVdb or Valsalva Ratio: n (%)<sup>1</sup> | 1 (4.5%) | 0 (0%) | 0 (0%) | — | — |
| **Sympathetic cholinergic variables (sudomotor)<sup>5</sup>** |  |  |  |  |  |
| Q-Sweat volume forearm | 0.52 ± 0.74 | — | — | — | — |
| Q-Sweat volume proximal leg | 0.70 ± 0.84 | — | — | — | — |
| Q-Sweat volume distal leg | 0.56 ± 0.60 | — | — | — | — |
| Q-Sweat volume foot | 0.50 ± 0.79 | — | — | — | — |
| Abnormal Q-Sweat: n (%) | 9 (64.3%) | — | — | — | — |
| Length dependent: n (%) | 4 (28.6%) | — | — | — | — |

Statistical tests were conducted based on data distribution normality. Parametric tests were done using a one-way analysis of variance (ANOVA) followed by Turkey HSD or Games-Howell<sup>a</sup> test based on equal variance adjusted for multiple comparisons. Non-parametric tests were done using a Kruskal-Wallis's<sup>b</sup> test followed by Dunn's post hoc test with Bonferroni adjustment for multiple comparisons. Statistical significance (\*) is shown for P-value < 0.05. One patient with IH and one control were excluded due to missing autonomic data. <sup>1</sup>data available for 22 participants with IH. <sup>2</sup>data available for 19 participants with IH. <sup>3</sup>data available for 17 participants with IH. <sup>4</sup>data available for 21 participants with IH. <sup>5</sup>data available for 14 participants with IH. Abbreviations: HUT - head-up tilt table testing; SBP – systolic blood pressure; DBP – diastolic blood pressure; HR – heart rate; Δ – change from baseline; HRVdb – heart rate variability during deep breathing; Q-Sweat – quantitative sudomotor axon reflex test; bpm – beats per minute; mmHg – millimeters of mercury.

**Figure 2.**
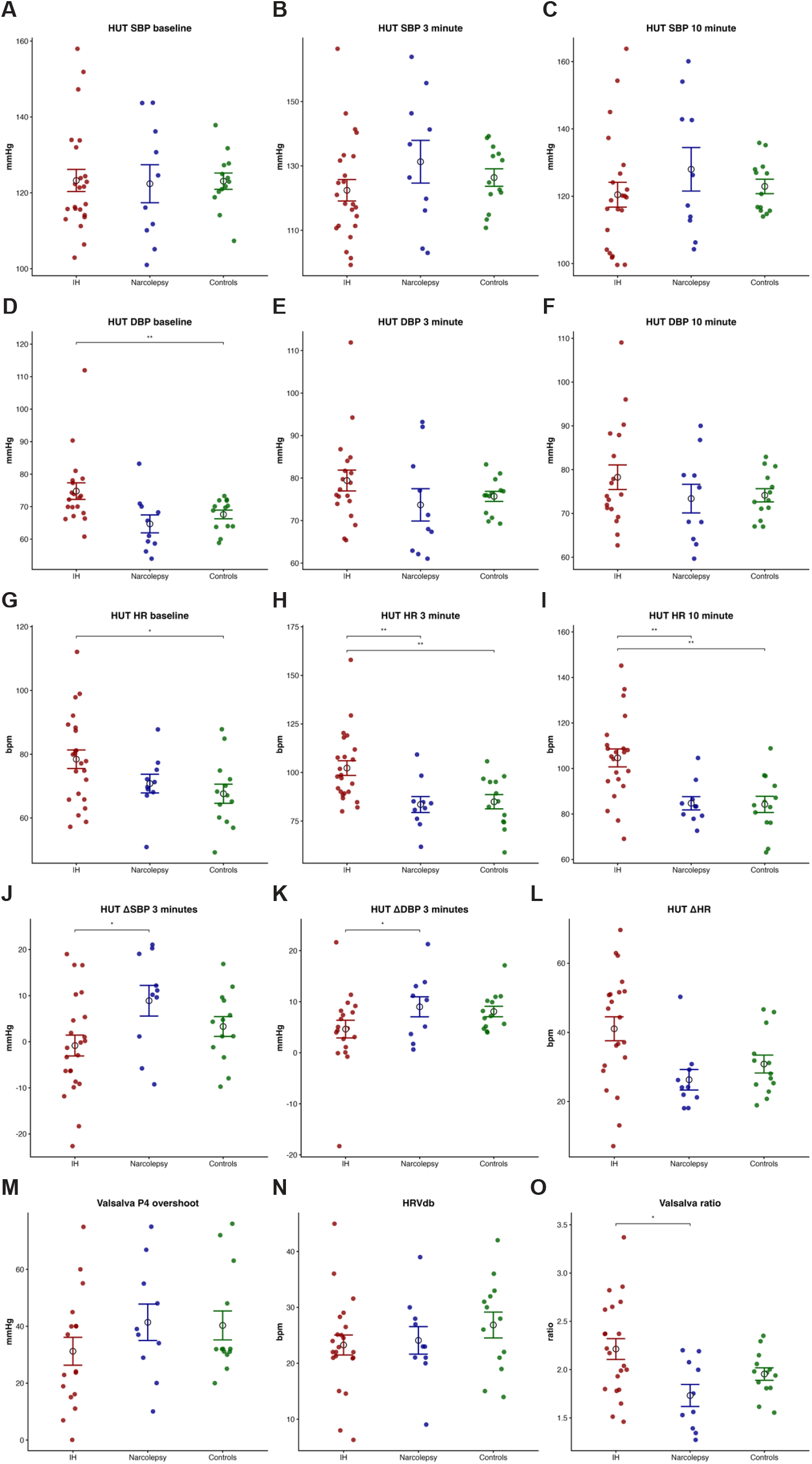
Autonomic reflex testing results across diagnostic groups. Panels A–C: Systolic blood pressure (SBP) at baseline, 3 minutes, and 10 minutes of head-up tilt (HUT). Panels D–F: Diastolic blood pressure (DBP) at baseline, 3 minutes, and 10 minutes. Panels G–I: Heart rate (HR) at baseline, 3 minutes, and 10 minutes of HUT. Panels J–K: ΔSBP and ΔDBP at 3 minutes of tilt. Panel L: Maximal ΔHR during HUT. Panel M: Valsalva P4 overshoot. Panel N: HRV during deep breathing (HRVdb). Panel O: Valsalva ratio. Each dot represents an individual participant; black bars indicate mean ± SEM. IH = idiopathic hypersomnia; NT1 = type-1 narcolepsy; HC = healthy controls.

Baseline SBP during HUT was similar across groups (Figure 2A–C), whereas DBP at baseline was significantly higher in IH compared to NT1 (p = 0.0077; Figure 2D). During HUT, IH participants had significantly greater HRs at 3 minutes compared to both NT1 and HC (Figure 2H), and this pattern persisted at 10 minutes (p < 0.001; Figure 2I). The HR increase from supine to peak during HUT (ΔHR) was also greater in IH than NT1 (Figure 2L), highlighting a marked orthostatic tachycardia in IH. In addition, orthostatic BP responses differentiated groups: ΔSBP at 3 minutes was significantly lower in IH compared with NT1 (Figure 2J). Abnormal HUT responses (orthostatic hypotension or exaggerated tachycardia by predefined consensus criteria) were observed in 69.6% of IH, whereas no NT1 or HC participants demonstrated an abnormal HUT response (Table 2).^16^

As a result, the greatest orthostatic HR and ΔHR increases were observed in IH, compared to NT1 and HCs (Figures 2G-L). Other sympathetic adrenergic and cardiovagal indices were largely preserved. Valsalva phase 4 BP overshoot (Figure 2M) and HRV during deep breathing (HRVdb; Figure 2N) did not differ among groups and only one IH participant had an abnormal cardiovagal value. The Valsalva ratio was significantly greater in IH compared to NT1, however mean vaules for both groups remained within the normal range (Figure 2O). 64.3% of IH participants had abnormal Q-Sweat sudomotor responses, suggestive of small-fiber neuropathy, with 28.6% demonstrating a length-dependent pattern and 35.7% demonstrating a non-length dependent pattern (Table 2). Due to slight differences in autonomic testing protocols across sites, sudomotor testing was not available for NT1 or HCs, precluding group comparisons.

Within IH, autonomic reflex profiles were broadly similar between those who reported long sleep (TST > 11 h) and those with TST ≤ 11 h (Supplementary Table 1; Supplementary Figure S1). Orthostatic HR and BP responses, HUT ΔHR, and cardiovagal indices (HRVdb and Valsalva ratio) did not differ significantly between subgroups. However, there was a trend toward more severe distal sudomotor impairment in the non–long⍰sleep subgroup, as Q⍰Sweat foot volumes were lower in IH with TST ≤ 11 h (0.1 ± 0.1 µL/5 min vs. 0.8 ± 1.0 µL/5 min, p = 0.033; Figure S1S), and abnormal QSART was numerically more frequent (100% vs. 37.5%, p = 0.052).

### Polysomnography and Sleep Architecture

PSG and MSLT findings for the full cohort are summarized in Table 3 and Supplementary Figure S2. Mean sleep onset latency on MSLT differed significantly across groups (p < 0.001), with NT1 demonstrating the shortest latency compared to IH and HCs. SOREMs were common in NT1 (4.0 ± 1.0 SOREMs) and rare/absent in IH and HCs (0.17 ± 0.39 and 0, respectively). Overnight PSG macroarchitecture (Table 3) also differed significantly among groups, with REM latency markedly prolonged in IH compared to both NT1 and HCs. Sleep efficiency was relatively high across groups (approximately 76–83%) and did not differ significantly. Total sleep time was also similar, averaging 6.8– 7 hours in all three groups.

**Table 3.** Polysomnography-derived sleep macroarchitecture parameters among patients with idiopathic hypersomnia (IH), narcolepsy, and healthy controls (HC).

| Sleep Variables | IH (n = 24)<br>Mean ± SD<br>Median (IQR) | Narcolepsy (n = 10)<br>Mean ± SD<br>Median (IQR) | Controls (n = 14)<br>Mean ± SD<br>Median (IQR) | P-value | Post-hoc |
| --- | --- | --- | --- | --- | --- |
| **Multiple Sleep Latency Test<sup>1</sup>** |  |  |  |  |  |
| MSLT (minutes) | 7.1 ± 3.5<br>6.2 (4.5) | 4.0 ± 1.2<br>3.9 (2.2) | 18.0 ± 2.0<br>18.0 (2.0) | <0.001* | — |
| Number of sleep-onset REM periods | 0.17 ± 0.39<br>0 (0) | 4.0 ± 1.0<br>4.0 (1.7) | 0<br>0 | <0.0001* | — |
| **Overnight Polysomnography** |  |  |  |  |  |
| Sleep-onset latency (minutes)<sup>2</sup> | 41.0 ± 79.6<br>19.7 (21.4) | 10.4 ± 18.2<br>0 (10.0) | 27.1 ± 39.2<br>12.0 (31.6) | 0.024*<sup>b</sup> | IH vs. NC |
| REM-sleep latency during (min)<sup>3</sup> | 154.8 ± 98.2<br>125.5 (102.5) | 59.9 ± 57.9<br>72.5 (80.7) | 64.6 ± 30.4<br>66.2 (19.0) | <0.01*<sup>b</sup> | IH vs. NC/HC |
| Sleep efficiency (%)<sup>4</sup> | 83.1 ± 18.9<br>87.8 (10.0) | 76.4 ± 10.6<br>75.0 (13.5) | 81.4 ± 14.2<br>85.2 (13.4) | 0.10<sup>b</sup> | — |
| TST (min)<sup>4</sup> | 408.8 ± 113.1<br>412.5 (85.0) | 415.4 ± 73.8<br>404.0 (87.5) | 386.1 ± 72.1<br>389.5 (62.2) | 0.51<sup>b</sup> | — |
| WASO (min)<sup>5</sup> | 38.4 ± 23.2<br>38.5 (26.9) | 117.1 ± 53.2<br>129.0 (89.2) | 66.4 ± 40.7<br>58.5 (20.1) | <0.0001*<sup>a</sup> | IH vs. NC |
| Stage N1 sleep (% TST)<sup>6</sup> | 9.6 ± 11.5<br>5.5 (7.3) | 17.9 ± 7.2<br>16.5 (5.0) | 5.4 ± 3.5<br>4.0 (2.5) | <0.001*<sup>b</sup> | IH vs. NC<br>NC vs. HC |
| Stage N2 sleep (% TST)<sup>6</sup> | 55.3 ± 12.8<br>52.8 (17.7) | 38.1 ± 9.2<br>39.0 (12.5) | 44.2 ± 7.5<br>45.4 (10.8) | <0.001* | IH vs. NC/HC |
| Stage SWS sleep (% TST)<sup>6</sup> | 16.6 ± 10.5<br>15.8 (14.0) | 20.3 ± 7.7<br>20.0 (11.0) | 24.0 ± 5.7<br>25.2 (4.3) | 0.058 | — |
| REM sleep (% TST)<sup>6</sup> | 17.7 ± 10.5<br>16.1 (15.9) | 23.9 ± 5.6<br>25.5 (9.5) | 26.3 ± 7.5<br>26.0 (11.1) | 0.012* | IH vs. HC |

Parametric tests were done using a one-way analysis of variance (ANOVA) followed by Turkey HSD or Games-Howell<sup>a</sup> test based on equal variance adjusted for multiple comparisons. Non-parametric tests
were done using a Kruskal-Wallis<sup>b</sup> test followed by Dunn's post hoc test with Bonferroni adjustment for multiple comparisons. Statistical significance (\*) is shown for P-value < 0.05. <sup>1</sup>data available for 23 participants with IH and 3 controls and only an independent T-test was conducted for IH vs. NC. <sup>2</sup>data available for 20 participants with IH. <sup>3</sup>data available for 19 participants with IH. <sup>4</sup>data available for 21 persons with IH. <sup>5</sup>data available for 18 participants with IH. <sup>6</sup>data available for 19 participants with IH. Abbreviations: TST – total sleep time; WASO – wake after sleep onset.

Stage distribution showed robust group differences (Table 3; Supplementary Figure S2). IH was characterized by an increased proportion of N2 sleep (≈55–60% of TST) and reduced REM sleep (≈17– 18%), whereas NT1 had more N1 sleep (≈18%) and less N2, and HCs had a more balanced distribution with higher N3 and REM percentages. WASO was substantially greater in NT1 than IH and REM% was less in IH relative to HCs. ROC analyses (Supplementary Figure S3) demonstrate that REM latency, WASO, N1%, and N2% discriminate IH from NT1 and controls with AUC values up to 0.92.

Within IH, sleep macroarchitecture was remarkably similar between those with and without long sleep (Supplementary Table 2). Age, MSLT latency, number of SOREMs, TST, sleep efficiency, WASO, and the proportions of N2, N3, and REM sleep did not differ significantly between IH participants with TST > 11 h and those with TST ≤ 11 h. While was a non⍰significant trend toward a higher N1 percentage in the long⍰sleep subgroup (13.4 ± 15.0% vs. 5.4 ± 2.9%; p = 0.094), the long⍰sleep phenotype did not show a distinct nocturnal macroarchitecture.

### Heart Rate Variability (HRV) During Overnight Polysomnography

In the subset with full PSG data (Supplementary Table S3), mean HR and key HRV indices across wake, N3, and REM (11 IH, 10 NT1, 14 HCs) are summarized in Table 4 and shown in Figure 3; ART results are in Supplementary Table 4 and sleep macroarchitecture in Supplementary Table S5.

**Table 4.**
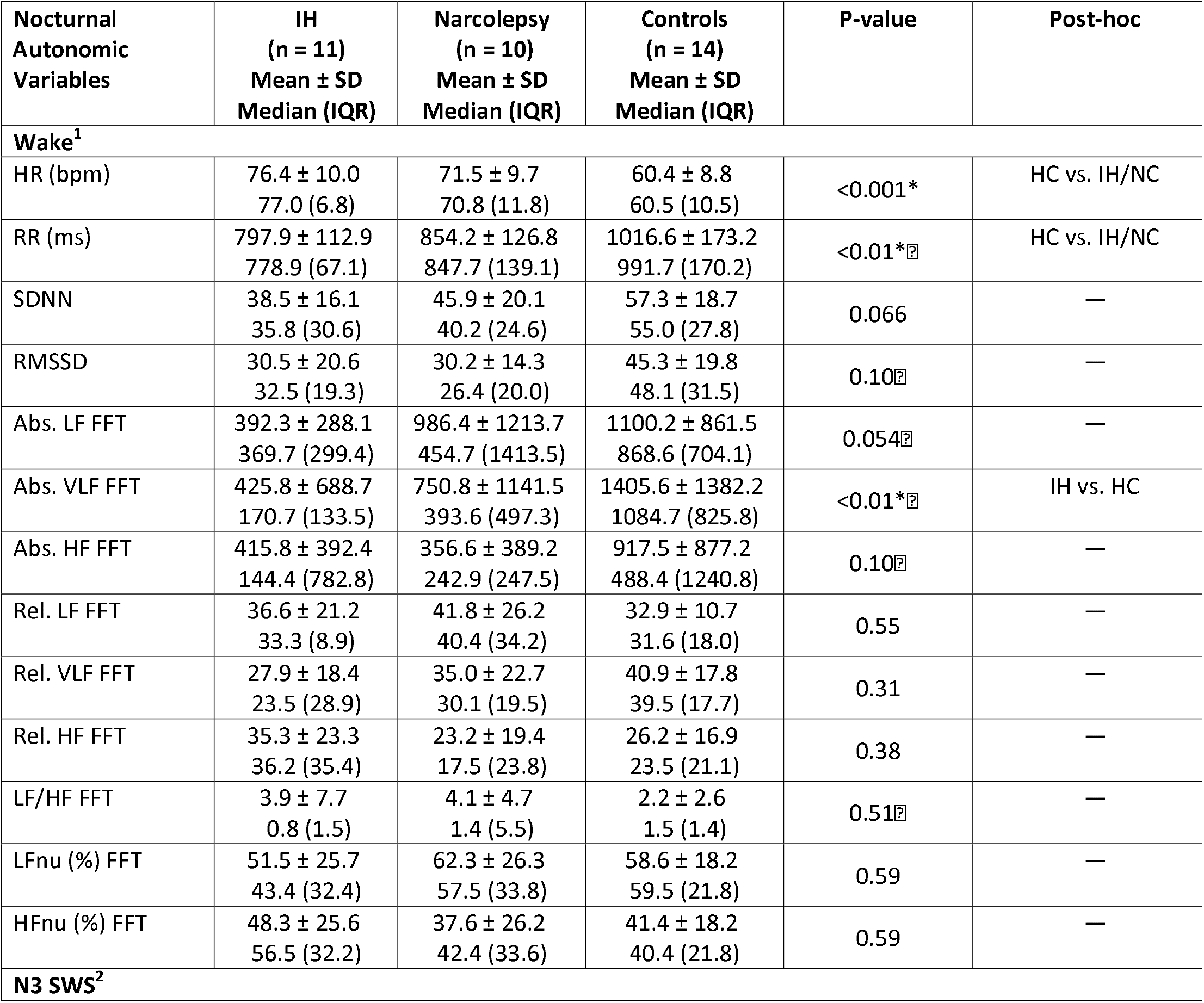
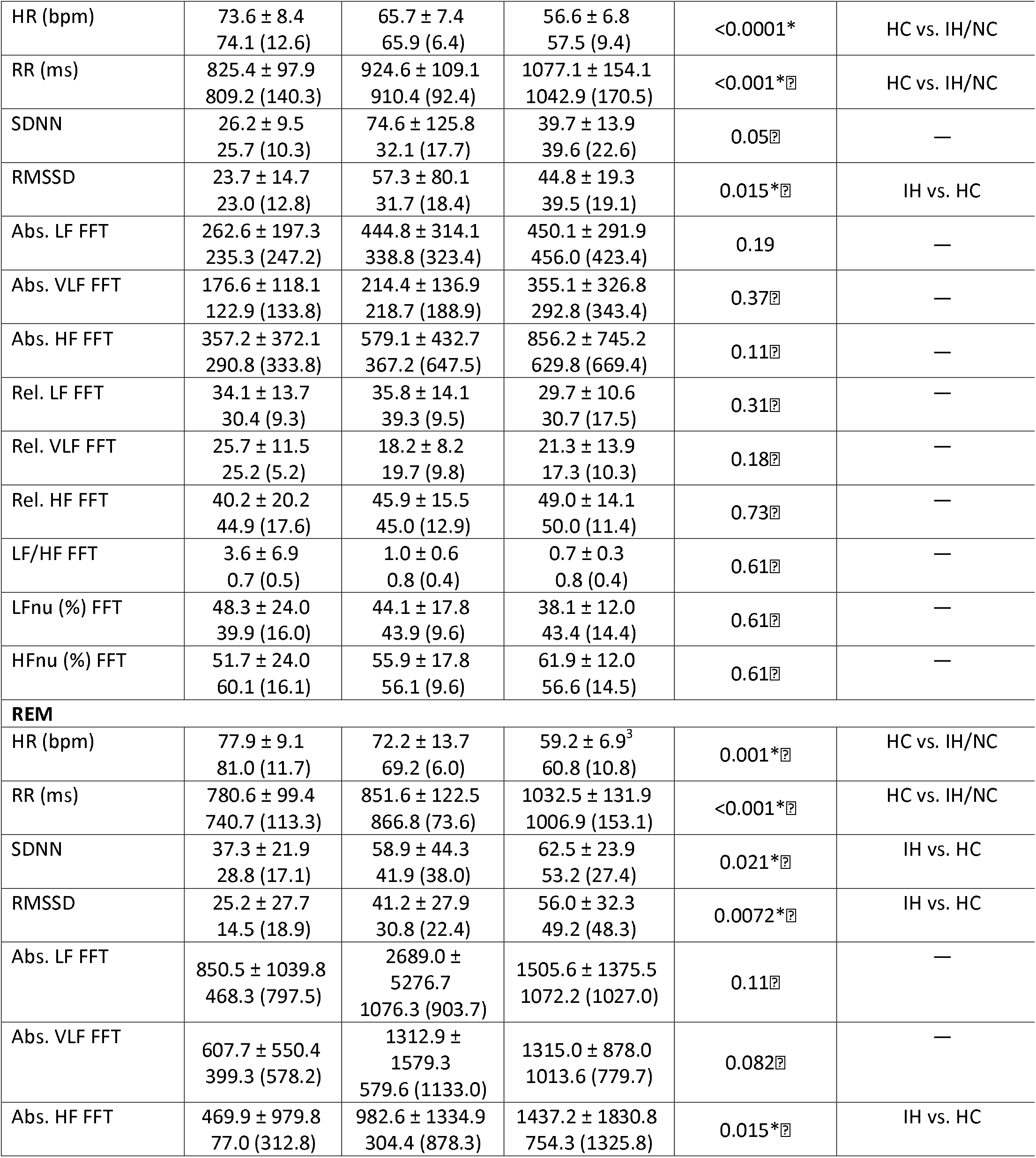
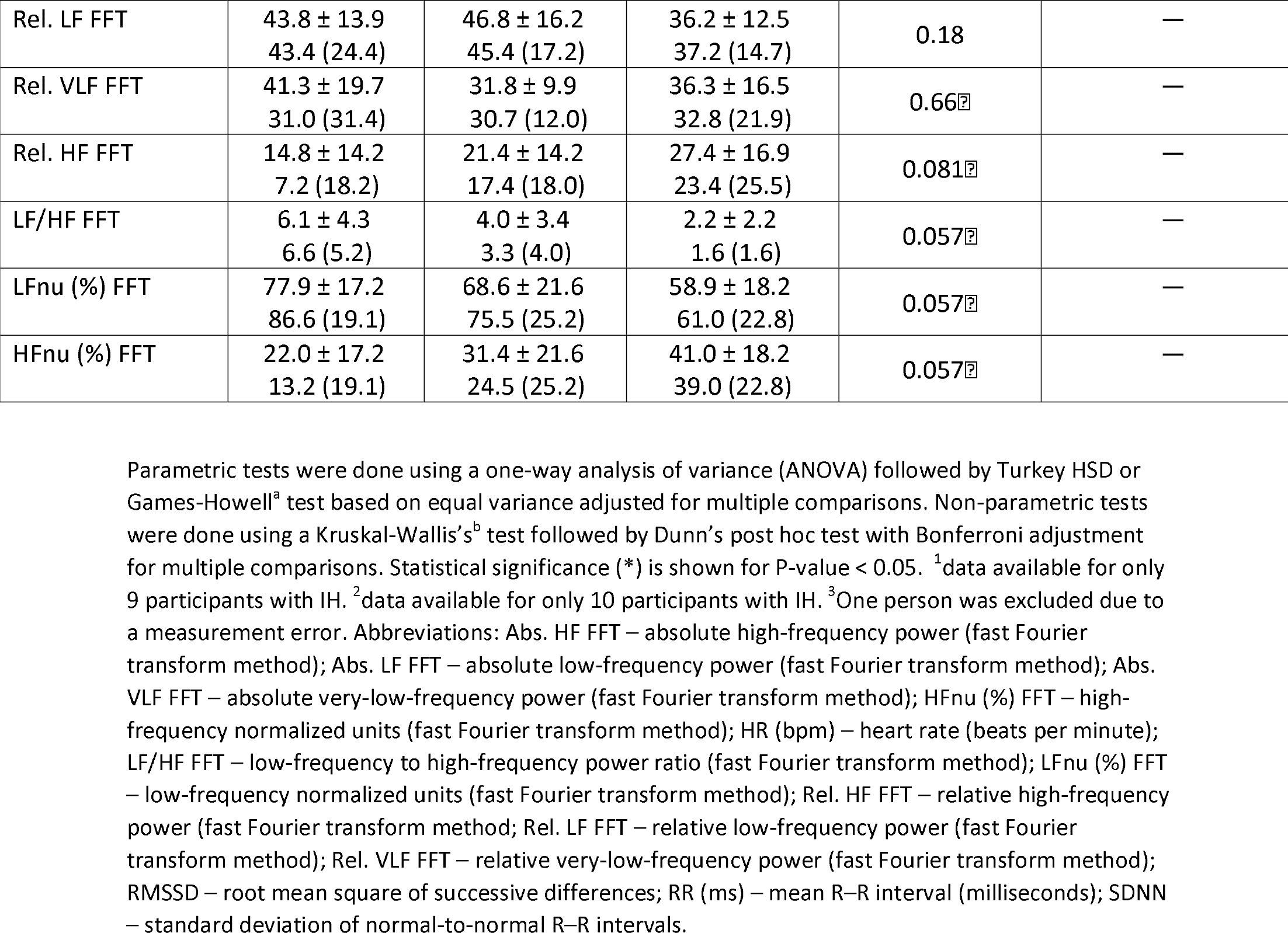
Polysomnography-derived sleep microarchitecture parameters among patients with idiopathic hypersomnia (IH), narcolepsy, and healthy controls (HC).

**Figure 3.**
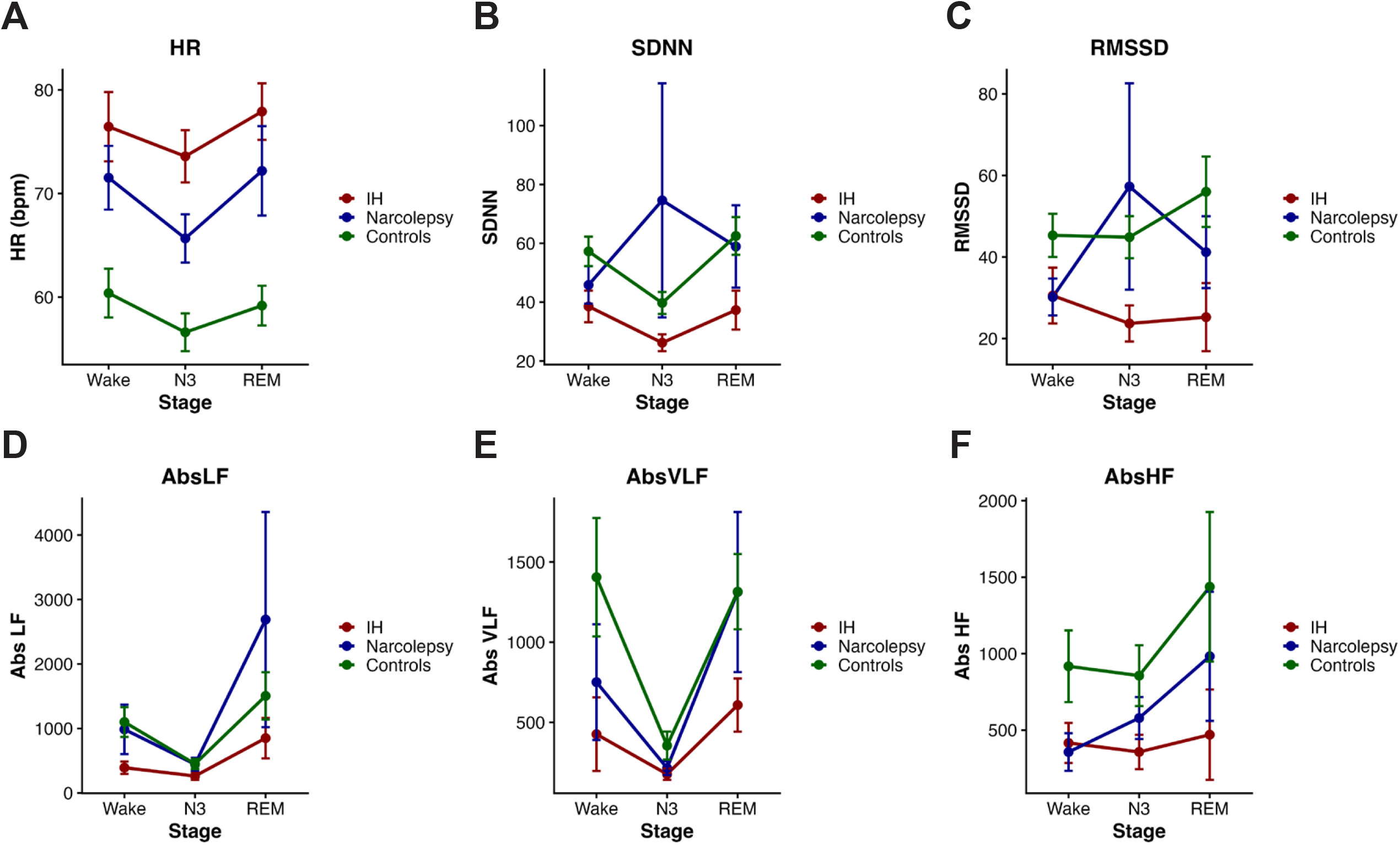
Heart rate and heart rate variability across vigilance states in idiopathic hypersomnia (IH), narcolepsy, and healthy controls. (A) Heart rate (HR, bpm). (B) SDNN (standard deviation of normal-to-normal RR intervals; overall HRV). (C) RMSSD (root mean square of successive RR interval differences; short-term/vagal HRV). (D) AbsLF (absolute low-frequency power). (E) AbsVLF (absolute very-low-frequency power). (F) AbsHF (absolute high-frequency power). Mean (± error bars) values are shown for Wake, N3 (slow-wave sleep), and REM sleep for each group (IH = red, narcolepsy = blue, controls = green).

Across all analyzed stages, both IH and NT1 participants had greater HRs (shorter RR intervals) than HCs, with the largest differences shown during N3 and REM (Table 4; Figure 3A). Time-domain HRV indices demonstrated reduced HRV and cardiovagal modulation in IH compared to controls, most evident during N3 and REM sleep; during REM, SDNN (Figure 3B) and RMSSD (Figure 3C) were both lower in IH than in healthy controls, with NT1 showing intermediate values, and similar reductions were observed during N3.

Frequency⍰domain measures further supported diminished variability and a shift in sympathovagal balance in IH (Table 4). In wake, IH participants demonstrated reduced very⍰low⍰frequency (VLF; Figure 3E)) power when compared to HCs, suggesting reduced slow⍰oscillatory components of HRV. During N3, autoregression (AR)⍰based microarchitecture analyses showed significantly lower absolute HF power in IH compared to HCs (Figure 3F), again consistent with reduced parasympathetic influence. In REM, both FFT⍰ and AR⍰based analyses revealed lower HF power and reduced total variability in IH relative to controls (Figure 3F), alongside lower VLF power (p ≈ 0.014–0.082). At the same time, REM⍰stage AR analyses demonstrated relatively higher LF contributions in IH, with correspondingly reduced HF percentages and a trend toward higher LF/HF ratios.

Taken together, the data (Table 3, Figure 4) demonstrates that IH is characterized by (1) higher HR across sleep stages, (2) reduced overall HRV, reflected by lower SDNN and lower absolute spectral power measures, (3) reduced cardiovagal modulation, reflected by lower RMSSD and HF power, and (4) a REM⍰specific pattern of relatively greater LF contribution with reduced HF contribution, consistent with sympathetic predominance or reduced parasympathetic buffering relative to controls. NT1 participants shared some features, including higher HR and shorter RR intervals, but reductions in HRV indices were generally less pronounced than IH. Stage⍰to⍰stage comparison heatmaps in Supplementary Figure S4 provide an additional visualization of how these HRV indices evolve across wake, N3, and REM in each group.

**Figure 4.**
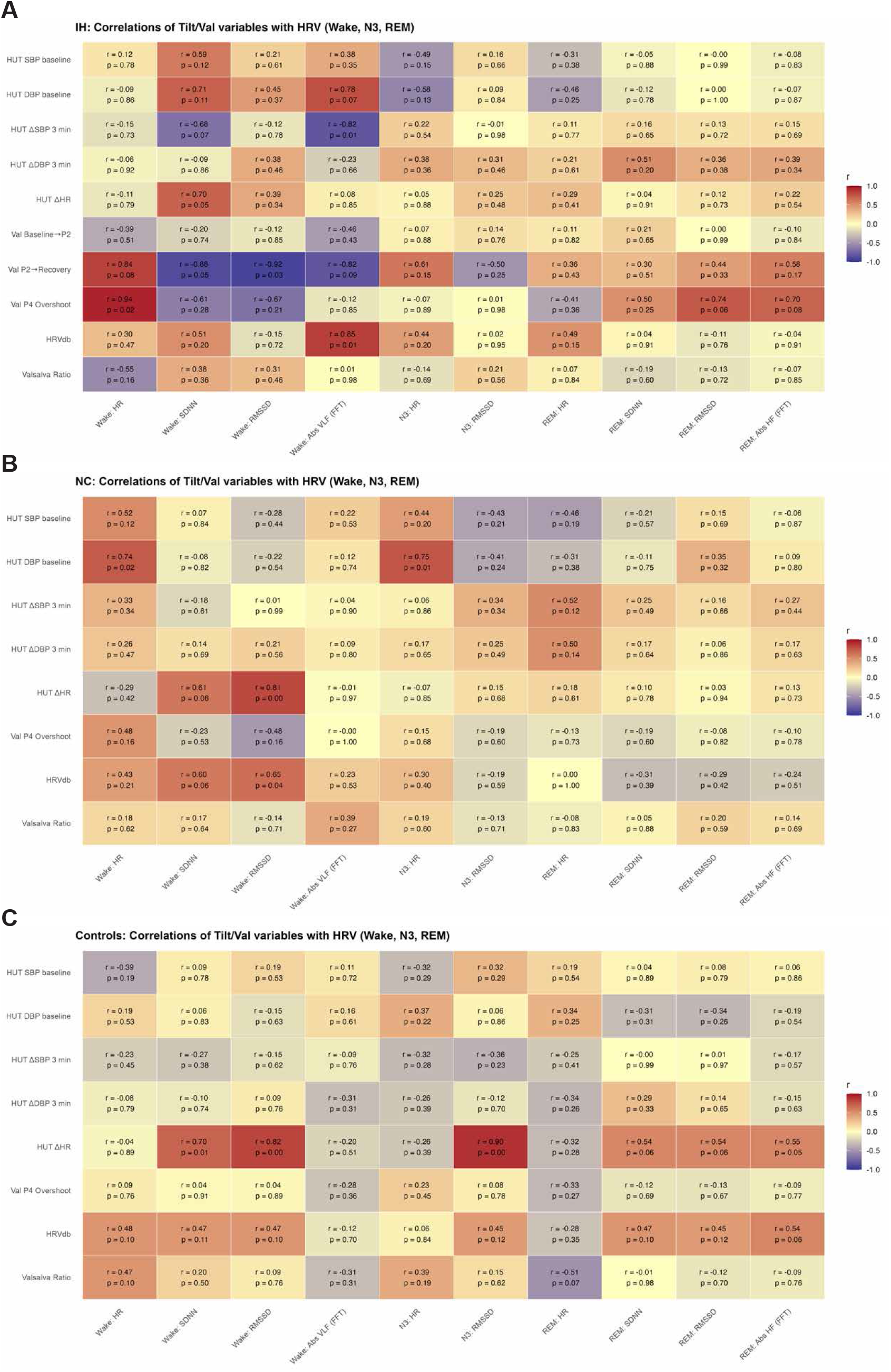
Group-specific correlations between daytime autonomic reflex indices and nocturnal cardiac measures. Heatmaps show Pearson correlation coefficients (r) linking diurnal autonomic testing metrics (rows; Valsalva adrenergic indices and head-up tilt [HUT] heart-rate response) with nocturnal heart rate (HR) and heart rate variability (HRV) measures during Wake, N3, and REM (columns). Panels depict (A) idiopathic hypersomnia (IH), (B) narcolepsy type 1 (NT1), and (C) healthy controls (HCs). Cell color indicates direction and magnitude of association (blue = positive; red = negative; deeper color = stronger correlation), and each cell is annotated with r and p values. Nocturnal HRV includes time-domain indices (SDNN, RMSSD) and frequency-domain indices (AbsVLF, AbsLF, AbsHF). Abbreviations: HR, heart rate; SDNN, standard deviation of normal-to-normal RR intervals; RMSSD, root mean square of successive RR interval differences; AbsVLF/AbsLF/AbsHF, absolute very-low/low/high frequency power; HUT ΔHR, change in HR with head-up tilt; P2, phase II; P4, phase IV.

### Correlations Between Diurnal Autonomic Reflex Testing and Nocturnal HRV

Correlation analyses linking diurnal autonomic reflex measures with nocturnal HRV demonstrated group-specific patterns (Figure 4). In IH, adrenergic Valsalva indices showed the strongest association with HRV: greater Valsalva phase 4 BP overshoot correlated with faster mean wake HRs (r = 0.94, p = 0.02) and phase 2-to-recovery magnitude correlated inversely with wake SDNN and RMSSD (r = -0.88, p = 0.05; r = -0.92, p = 0.03), suggesting that exaggerated Valsalva BP overshoot (a sign of sympathetic hyperactivity) is coupled with reduced nocturnal variability and vagal modulation. Orthostatic tachycardia (HUT ΔHR), however, showed only modest and inconsistent correlations with sleep-stage HRV in the IH group. NT1 participants demonstrated strong positive correlations between ΔHR and wake RMSSD (r = 0.81, p < 0.001) and in HCs a similar relationship was observed between ΔHR and N3 RMSSD (r = 0.90, p < 0.001). These patterns, visualized in Figure 3, suggest that in NT1 and HCs, diurnal orthostatic responses and nocturnal vagal modulation remain more tightly coordinated, whereas in IH this coupling is disrupted and replaced by a stronger association between exaggerated adrenergic responses (Valsalva overshoot) and reduced nocturnal variability.

## DISCUSSION

In this study we combined standardized diurnal autonomic reflex testing with nocturnal HRV analyses to characterize autonomic dysfunction in IH compared with NT1 and HC. Several findings emerged. First, IH was associated with marked orthostatic tachycardia on head-up tilt and frequent sudomotor abnormalities, consistent with clinically significant cardiovascular and peripheral autonomic dysfunction. In our cohort, a majority of those with IH met criteria for exaggerated postural tachycardia on tilt (ΔHR ≥ 30 bpm), and most of these were accompanied by symptoms of orthostatic intolerance; however, because symptom chronicity was not uniformly available, we describe these findings as orthostatic tachycardia rather than uniformly labeling them postural tachycardia syndrome (POTS). Second, during sleep, both IH and NT1 participants exhibited evidence of reduced cardiovagal modulation and greater HRs relative to controls, with IH participants exhibiting the most pronounced reduction in overall HRV and cardiovagal function, particularly during REM sleep. Third, REM-related autonomic control appeared especially abnormal in IH, with greater HRs and lower SDNN, RMSSD, and HF power, indicating a shift toward sympathetic predominance and reduced parasympathetic/vagal buffering during a physiologically unstable sleep stage. Notably, diurnal cardiovagal reflex indices (HRVdb and Valsalva ratio) were relatively preserved, suggesting a state-dependent phenotype in which nocturnal cardiovagal modulation is reduced despite intact diurnal cardiovagal reflex responses. Together, these findings provide convergent diurnal and nocturnal evidence that IH is accompanied by substantial autonomic dysregulation that is at least as great as, and in some respects exceeds, that seen in NT1.

HRV is a noninvasive measure of cardiac autonomic modulation derived from beat-to-beat (NN/RR) interval variation. In this study, we focused on time-domain indices (SDNN, RMSSD) and frequency-domain indices (HF and LF power). RMSSD and HF power (0.15–0.40 Hz) predominantly reflect respiratory sinus arrhythmia and cardiovagal modulation, whereas SDNN reflects overall variability over the analyzed epoch and LF power reflects mixed influences that include baroreflex-mediated autonomic activity.^8–11^ We interpreted nocturnal HRV using complementary indices: RMSSD and HF power as markers of cardiovagal modulation, SDNN as an index of overall variability, and LF power as a mixed measure influenced by baroreflex mechanisms; consistent with prior work, we did not interpret LF/HF as a simple index of “sympathovagal balance.”^8–11^

HRV exhibits circadian periodicity and undergoes significant changes during the transition from wake to sleep and across sleep stages.^11^ Previous research has demonstrated that vagal HRV measures (such as HF and RMSSD) increase in activity during sleep, accompanied by a reduction in heart rate, indicating stronger parasympathetic modulation during nocturnal sleep compared to daytime measures.^17,18^ Additionally,, autonomic control differs between NREM and REM sleep: NREM is associated with lower sympathetic activity, reduced cardiac output, and lower BP, whereas REM sleep is characterized by greater sympathetic activity and greater cardiovascular lability than quiet wakefulness.^19–22^ A large body of work has demonstrated that reduced overall HRV and reduced vagal indices are associated with worse cardiovascular outcomes in the general population as well as several disease states.^11^

HRV abnormalities have been well described in NT1 and appear to arise from orexin neuron loss, which disrupts descending hypothalamic projections to autonomic control centers and contributes to altered heart rate and blood pressure regulation.^23^ In IH, by contrast, objective autonomic data have been limited. One prior HRV study suggested increased parasympathetic activity during wake and sleep in IH, whereas questionnaire-based work has consistently highlighted a high burden of autonomic symptoms.^4^ Our data extend this literature by integrating diurnal autonomic reflex testing with stage-specific nocturnal HRV in a well-characterized IH cohort, and by directly comparing IH with NT1 and HC.

Our autonomic reflex data extend prior symptom-based work showing a high prevalence of orthostatic intolerance, palpitations, and other autonomic complaints in IH.^3^ We now demonstrate that these symptoms are accompanied by objective abnormalities on standardized autonomic reflex testing. IH participants exhibited significantly greater ΔHR during head⍰up tilt than both NT1 and HC, with approximately 70% of IH participants demonstrating an exaggerated postural tachycardia. Sudomotor testing further revealed that nearly two⍰thirds of IH participants who underwent testing had abnormal sudomotor responses, with most demonstrating a non-length dependent pattern, a more typical pattern of immune-mediated small fiber neuropathy.^18^ It should be noted that we did not have detailed neurological exams on all participants, nor could we corroborate abnormal testing results with symptoms of neuropathy in our cohort. Although we lacked sudomotor data in NT1 and HC, these rates of abnormal sweating responses in IH suggests that peripheral autonomic involvement may co-occur with prominent abnormalities of cardiovascular autonomic control (orthostatic tachycardia) in this disorder. Standard bedside cardiovagal indices (HRV during deep breathing and Valsalva ratio) were largely within normal limits, with only a minority of IH patients demonstrating diurnal cardiovagal abnormalities.

Across wake, SWS, and REM sleep, both IH and NT1 exhibited higher HRs and shorter RR intervals compared to controls, consistent with heightened sympathetic drive or reduced cardiovagal drive. However, IH participants exhibited a more pronounced reduction in overall HRV and cardiovagal indices relative to HC, most prominently during REM sleep, where SDNN, RMSSD, and HF power were all significantly lower. Given that RMSSD and HF primarily reflect respiratory sinus arrhythmia and short-latency vagal influences, these findings suggest reduced cardiovagal function in IH compared to controls during a physiologically vulnerable sleep stage.

The REM findings are particularly noteworthy. In healthy individuals, REM sleep is characterized by surges in sympathetic activity and greater cardiovascular instability compared to NREM sleep, however robust vagal modulation can buffer these fluctuations.^21^ In our cohort, IH participants not only exhibited greater HRs during REM than controls, but also exhibited reduced parasympathetic modulation, suggesting a double hit of increased sympathetic output and impaired vagal braking. This combination is classically associated with higher cardiovascular risk in general populations and other sleep disorders.^11^ Although our study was not designed to assess clinical outcomes, the pattern raises the possibility that IH may carry cardiovascular consequences beyond sleepiness and quality⍰of⍰life impairment. In fact, a recent claims-based analysis demonstrated that diagnoses of IH were associated with a higher odds of prevalent cardiovascular conditions or events than matched controls.^19^

Our results differ in important ways from another HRV study that found higher HF power and HFnu with a lower LF/HF ratio in IH across sleep cycles, consistent with a relative increase in parasympathetic activity and an exaggerated sympathetic response to arousals.^5^ Several methodological and sample differences may explain this apparent discrepancy. This study employed 24⍰hour ambulatory PSG in a smaller IH cohort without comorbid sleep disorders, and HRV was averaged across entire sleep cycles, including arousals and respiratory events. In contrast, we analyzed carefully selected artifact⍰free 5⍰minute segments of stable SWS and REM sleep and directly compared IH to both NT1 and HC. Differences in disease duration, sex distribution (our IH cohort was predominantly female), medication exposure, and autonomic symptom burden may also contribute. Rather than contradicting these authors’ findings, our results are likely to reflect both analytic perspective and biologic heterogeneity within IH. The authors also quantified HRV across averaged sleep cycles, incorporating multiple phases and transitions (and thus capturing arousal-related dynamics), and reported relatively greater parasympathetic indices with heightened autonomic responses around arousals. In contrast, we used a “magnifying lens” approach, analyzing artifact-free 5-minute segments within stable N3 and REM sleep, intended to estimate stage-specific baseline autonomic modulation. From this perspective, our cohort, enriched for patients with marked orthostatic tachycardia, showed lower cardiovagal indices during sleep, particularly in REM. Together, these data support the possibility that IH includes distinct autonomic phenotypes: some patients may exhibit higher baseline vagal tone with exaggerated reactivity to arousals, whereas others demonstrate reduced cardiovagal modulation and higher heart rate during stable sleep.

The NT1 HRV phenotype observed here is broadly consistent with prior work once methodological differences are considered. Studies have demonstrated that that hypocretin-deficient narcolepsy with cataplexy is characterized by consistently elevated HR across wake and sleep, with largely preserved HRV indices when sleep fragmentation and respiratory events are carefully controlled.^24^ Other studies similarly found that narcoleptic patients show normal stage-dependent variations in HRV, with lower HF and higher LF/HF during wakefulness before sleep and a reciprocal pattern across NREM and REM, without evidence of primary cardiac ANS failure.^25^ Against this backdrop, the more severe REM-related vagal reduction we observed in IH underscores that IH cannot be assumed to share the same autonomic physiology as NT1 despite overlapping clinical features.

The mechanisms underlying autonomic dysfunction in IH are likely multifactorial and may differ from those in NT1. In NT1, loss of hypocretin/orexin neurons disrupts projections from the lateral hypothalamus to autonomic control centers in the brainstem and spinal cord, contributing to altered sympathetic and parasympathetic outflow, non⍰dipping blood pressure profiles, and impaired HRV. In IH, hypocretin levels are typically normal, prompting consideration of alternative neuromodulatory, immune and peripheral mechanisms.^22^ Sforza and colleagues proposed a role for the histaminergic system based on reduced cerebrospinal fluid histamine in IH and narcolepsy without cataplexy, and on the known wake⍰promoting and autonomic effects of histamine, particularly via H3 receptor modulation.^5^ Histaminergic neurons in the tuberomammillary nucleus project broadly to cortical, hypothalamic, and brainstem autonomic nuclei, and interact closely with the orexin system.^23^ Perturbations in histamine–orexin signaling could therefore contribute to both excessive sleepiness and impaired baroreflex and cardiovagal function in central hypersomnias when orexin concentrations remain within the range of normal.

Our observation of reduced overall HRV and cardiovagal indices in IH, together with orthostatic tachycardia and frequent sudomotor deficits, is most consistent with a primary disturbance of autonomic regulation: central “set-points” appear shifted toward chronically higher sympathetic drive with weaker parasympathetic buffering. That pattern alone could explain why IH patients show both exaggerated HR responses to upright tilt and blunted vagal modulation during REM sleep. Deconditioning then sits on top of this as a plausible amplifier rather than a sole explanation. Studies have demonstrated that reduced physical fitness is strongly and independently associated with orthostatic intolerance.^26^ Patients with IH commonly spend long periods in bed, experience persistent fatigue, and reduce their day-to-day activity, making them particularly vulnerable to this deconditioning loop. At present it is not clear how much of the autonomic profile we observed reflects a primary feature of IH pathophysiology and how much reflects secondary adaptations to years of hypersomnolence and activity restriction; longitudinal and mechanistic studies will be needed to explore these components.

From a clinical perspective, our findings emphasize that IH should be considered a disorder with significant autonomic involvement, not solely a condition of excessive sleepiness. Orthostatic tachycardia of the magnitude observed here is likely to be symptomatic and may contribute to fatigue, brain fog, and exercise intolerance commonly reported by IH patients. Routine screening for orthostatic intolerance and other autonomic symptoms, followed by targeted testing (active stand testing or more detailed autonomic testing), may help identify patients who could benefit from targeted treatment.

Strengths of this study include the multimodal assessment of autonomic function, combining tilt, Valsalva, HRDB, Q-Sweat, and nocturnal HRV parameters; the inclusion of both NT1 and HC comparison groups, which allowed us to distinguish IH-specific autonomic features from those shared across central hypersomnias; and the use of rigorously selected artifact-free sleep segments for HRV analysis. Our sample also reflects real⍰world IH practice, with a high proportion of female participants and frequent long⍰sleep phenotypes.

Several limitations should be acknowledged. The overall sample size was modest, particularly in the NT1 group, which may have limited power to detect more subtle group differences or interactions between sleep stage and diagnosis. The retrospective designs preclude inference about causality or temporal evolution of autonomic dysfunction. Although major confounders such as untreated sleep apnea and known autonomic disorders were excluded, residual medication effects cannot be fully ruled out. Medications were held prior to testing in both cohorts, but baseline use of sleep- and autonomic-active medications differed across groups and longer-term effects on heart rate, HRV, or sleep architecture cannot be entirely excluded. Additionally, comorbid medical or psychiatric conditions and deconditioning may have contributed and could not be fully excluded. Differences in BMI across groups, particularly higher BMI in NT1, may have influenced some autonomic measures. Sudomotor data were not available for NT1 or control participants, preventing direct group comparisons for Q-Sweat and limiting our ability to determine whether sudomotor abnormalities are specific to IH or shared across central hypersomnias.

In summary, this study provides convergent diurnal and nocturnal evidence that IH is associated with significant autonomic dysfunction. IH patients exhibited pronounced diurnal orthostatic tachycardia, frequent sudomotor abnormalities, reduced overall nocturnal HRV, and diminished nocturnal cardiovagal modulation, particularly during REM sleep, where higher heart rates and lower SDNN, RMSSD, and HF power indicate a shift toward sympathetic predominance relative to both NT1 and healthy controls. These findings extend prior symptom-based reports of autonomic dysfunction in IH and suggest that autonomic disturbances may be an integral feature of the disorder rather than an incidental comorbidity. Future work should aim to replicate these results in larger cohorts to define autonomic phenotypes within IH using integrated symptoms, reflex, sudomotor and HRV data, and determine whether targeted treatment of autonomic dysfunction can improve symptoms, normalize HRV and reduce long-term cardiovascular risk.

## Data Availability

The data that support the findings of this study are available on request from the corresponding author. The data are not publicly available due to privacy or ethical restrictions.

## Disclosure statement

MGM has received research support from Embr Wave, Biohaven Pharmaceuticals, Argenx Pharmaceuticals, Dysautonomia International and the National Institutes of Health; consulting fees from Jazz Pharmaceuticals, 2nd MD, Infinite MD, and Guidepoint LLC; and royalties from Elsevier. All other authors have nothing to disclose.

## Notes

### Funding Statement

This study did not receive any funding

### Author Declarations

All subjects gave written informed consent, and the study was approved by the local Institutional Review Boards of Stanford University and the University of Bologna.

