## Supplementary for "Nocturnal and Diurnal Measures of Autonomic Function in Idiopathic Hypersomnia and Type 1 Narcolepsy"

**Supplementary Table 1. Summary of autonomic function testing results of patients with idiopathic hypersomnia (IH) stratified by subjective reporting of total sleep time > 11 hour**. Statistical tests were conducted using independent t-tests or Mann–Whitney U[a] test based on data distribution normality. Chi-square test conducted for categorical variables. Statistical significance (*) is shown for P-value < 0.05. [1]data available for 13 persons with IH (TST > 11 hours). [2]data available for 12 persons with IH (TST > 11 hours). [3]data available for 12 persons with IH (TST > 11 hours) and 7 persons with IH (TST < 11 hours). [4]data available for 11 persons with IH (TST > 11 hours) and 7 persons with IH (TST < 11 hours). [5]data available for 13 persons with IH (TST > 11 hours) and 9 persons with IH (TST < 11 hours). [6]data available for 9 persons with IH (TST > 11 hours) and 8 persons with IH (TST < 11 hours). [7]data available for 11 persons with IH (TST > 11 hours). [8]data available for 8 persons with IH (TST > 11 hours) and 6 persons with IH (TST < 11 hours). *Abbreviations*: HUT – head-up tilt table testing; SBP – systolic blood pressure; DBP – diastolic blood pressure; HR – heart rate; Δ – change from baseline; HRVdb – heart rate variability during deep breathing; QSART – quantitative sudomotor axon reflex test; bpm – beats per minute; mmHg – millimeters of mercury.

| **Variables** | **IH (TST > 11 hours) (n = 14)** | **IH (TST ≤ 11 hours) (n = 10)** | **P value** |
|---|---|---|---|
| **Sympathetic adrenergic BP and HR variables** | | | |
| HUT SBP (mmHg): | | | |
| Baseline[1] | 124.6 ± 14.8 | 121.5 ± 13.4 | 0.85[a] |
| 3 Min[1] | 121.4 ± 12.6 | 123.8 ± 20.2 | 0.75 |
| 10 Min[2] | 119.7 ± 15.5 | 121.4 ± 20.1 | 0.83 |
| HUT DBP (mmHg): | | | |
| Baseline[3] | 75.2 ± 12.8 | 74.1 ± 8.3 | 0.93[a] |
| 3 Min[3] | 78.4 ± 7.8 | 81.1 ± 15.0 | 0.67 |
| 10 Min[4] | 78.4 ± 7.8 | 78.7 ± 15.8 | 0.92 |
| HUT HR (bpm): | | | |
| Baseline[1] | 80.0 ± 15.3 | 76.4 ± 12.5 | 0.54 |
| 3 Min[1] | 105.5 ± 21.7 | 98.1 ± 11.7 | 0.31 |
| 10 Min[2] | 110.2 ± 18.2 | 98.0 ± 17.3 | 0.12 |
| HUT ΔSBP 3 minutes (mmHg)[1] | −3.2 ± 8.6 | 2.3 ± 13.0 | 0.26 |
| HUT ΔDBP 3 minutes (mmHg)[3] | 3.2 ± 7.7 | 7.0 ± 7.3 | 0.67[a] |
| HUT ΔHR[5] | 43.7 ± 16.2 | 37.2 ± 16.5 | 0.38 |
| Abnormal HUT: n (%) | 10 (76.9%) | 7 (70.0%) | 1.0 |
| Valsava baseline to P2 min (mmHg)[6] | −23.5 ± 35.3 | −1.0 ± 40.1 | 0.19[a] |
| Valsava P2 recovery (mmHg)[6] | 7.4 ± 9.8 | 15.9 ± 44.3 | 0.47[a] |
| Valsava P4 overshoot (mmHg)[6] | 38.0 ± 20.5 | 23.6 ± 18.2 | 0.15 |
| **Cardiovagal HR variables** | | | |
| HRVdb (bpm)[2] | 22.1 ± 10.2 | 24.7 ± 5.7 | 0.46 |
| Valsalva Ratio[7] | 2.3 ± 0.5 | 2.1 ± 0.4 | 0.45 |
| Abnormal HRVdb or Valsalva Maneuver: n (%) | 1 (8.3%) | 0 (0%) | 1.0 |
| **Sympathetic cholinergic dysfunction (sudmotor)[8]** | | | |
| QSART volume forearm | 0.8 ± 0.9 | 0.2 ± 0.2 | 0.061[a] |
| QSART volume proximal leg | 1.0 ± 1.0 | 0.3 ± 0.2 | 10 |

| | | | |
|---|---|---|---|
| QSART volume distal leg | 0.8 ± 0.7 | 0.3 ± 0.2 | 0.073 |
| QSART volume foot | 0.8 ± 1.0 | 0.1 ± 0.1 | 0.033*[a] |
| Abnormal QSART: n (%) | 3 (37.5%) | 6 (100%) | 0.052 |
| Length dependent: n (%) | 1 (14.3%) | 3 (50.0%) | 0.22 |

**Supplementary Table 2. Polysomnography-derived sleep macroarchitecture parameters among patients with idiopathic hypersomnia (IH) stratified by subjective reporting of total sleep time > 11 hour**. Statistical tests were conducted using independent t-tests or Mann–Whitney U[a] test based on data distribution normality. Chi-square test conducted for categorical variables. Statistical significance (*) is shown for P-value < 0.05. [1]data available for 11 persons with IH (TST > 11 hours) and 9 persons with IH (TST < 11 hours). [2]data available for 11 persons with IH (TST > 11 hours) and 10 persons with IH (TST < 11 hours). [3]data available for 9 persons with IH (TST > 11 hours) and 9 persons with IH (TST < 11 hours). [4]data available for 10 persons with IH (TST > 11 hours) and 9 persons with IH (TST < 11 hours)

| Variables | IH (TST > 11 hours) (n = 13) | IH (TST ≤ 11 hours) (n = 10) | P-value |
|---|---|---|---|
| **Mean Sleep Onset Latency Testing** | | | |
| MSLT (minutes) | 8.1 ± 4.2<br>7.1 (6.9) | 5.8 ± 2.1<br>5.4 (3.4) | 0.10 |
| Number of sleep-onset REM periods | 0.15 ± 0.38<br>0 (0) | 0.20 ± 0.42<br>0.0 (0.0) | 0.81[a] |
| **Overnight Polysomnography** | | | |
| Sleep-onset latency (minutes)[1] | 56.4 ± 105.6<br>25.7 (19.8) | 22.1 ± 19.3<br>15.0 (6.7) | 0.36[a] |
| REM-sleep latency during (min)[2] | 148.4 ± 95.8<br>123.2 (111.5) | 161.9 ± 106.1<br>125.5 (74.0) | 0.65[a] |
| Sleep efficiency (%)[1] | 80.2 ± 25.7<br>89.8 (11.9) | 86.4 ± 6.0<br>87.0 (4.7) | 0.78[a] |
| TST (min)[1] | 417.5 ± 151.3<br>430.0 (86.6) | 399.1 ± 53.0<br>401.0 (62.7) | 0.27[a] |
| WASO (min)[3] | 39.3 ± 29.7<br>25.0 (27.0) | 37.4 ± 16.2<br>43.5 (17.5) | 0.87 |
| Stage N1 sleep (% TST)[4] | 13.4 ± 15.0<br>8.6 (8.2) | 5.4 ± 2.9<br>4.6 (5.0) | 0.094[a] |
| Stage N2 sleep (% TST)[4] | 52.7 ± 13.8<br>48.9 (23.2) | 58.1 ± 11.6<br>54.7 (7.9) | 0.38 |
| Stage N3 sleep (% TST)[4] | 12.8 ± 9.9<br>13.1 (16.1) | 20.8 ± 9.9<br>21.0 (11.6) | 0.10 |
| REM sleep (% TST)[4] | 21.0 ± 11.2<br>23.3 (13.5) | 14.1 ± 8.9<br>13.1 (5.0) | 0.16 |

**Supplementary Table 3. Demographic and clinical characteristics of patients with idiopathic hypersomnia (IH), narcolepsy, and healthy controls who underwent polysomnography.** Statistical tests were done using a one-way analysis of variance (ANOVA) or chi-square test for categorical variables. Statistical significance (*) is shown for P-value < 0.05. *Abbreviations:* BMI – body mass index; SD – standard deviation; IQR – interquartile range.

| Variables | IH (n = 11)<br>Mean ± SD<br>Median (IQR) | Narcolepsy (n = 10)<br>Mean ± SD<br>Median (IQR) | Controls (n = 13)<br>Mean ± SD<br>Median (IQR) | P-value |
|---|---|---|---|---|
| Age (years) | 32.5 ± 7.7<br>32.0 (5.5) | 37.5 ± 7.6<br>35.5 (8.5) | 37.7 ± 10.4<br>33.0 (14.0) | 0.37 |
| Female/male (%) | 11 (100%) | 1 (10%) | 6/7 (46.1%) | <0.001* |
| BMI (kg/m$^2$) | 24.3 ± 4.8<br>24.9 (4.1) | 28.1 ± 3.6<br>27.8 (3.5) | 23.2 ± 4.6<br>22.8 (5.5) | 0.035* |
| Disease onset age | 30.5 ± 7.2<br>29.0 (5.5) | 13.6 ± 5.6<br>13.5 (6.8) | — | — |
| Disease duration (at autonomic testing) | 3.7 ± 3.2<br>23.9 (10.5) | 23.9 ± 10.5<br>21.0 (14.5) | — | — |

**Supplementary Table 4. Summary of autonomic function testing results in patients with idiopathic hypersomnia (IH), narcolepsy (NC), and healthy controls (HC) who underwent polysomnography**. Parametric tests were done using a one-way analysis of variance (ANOVA) followed by Turkey HSD or Games-Howell[a] test based on equal variance adjusted for multiple comparisons. Non-parametric tests were done using a Kruskal-Wallis's[b] test followed by Dunn's post hoc test with Bonferroni adjustment for multiple comparisons. Statistical significance (*) is shown for P-value < 0.05. [1]data available for 10 persons with IH. [2]data available for 8 persons with IH. [3]data available for 7 persons with IH. [4]data available for 9 persons with IH. *Abbreviations*: HUT – head-up tilt table testing; SBP – systolic blood pressure; DBP – diastolic blood pressure; HR – heart rate; Δ – change from baseline; HRVdb – heart rate variability during deep breathing; QSART – quantitative sudomotor axon reflex test; bpm – beats per minute; mmHg – millimeters of mercury.

| Variables | IH (n = 11) | Narcolepsy (n = 10) | Controls (n = 13) | P value | Post-hoc |
|---|---|---|---|---|---|
| **Sympathetic adrenergic BP and HR variables** | | | | | |
| HUT SBP (bpm)[1]:<br>Baseline<br>3 Min<br>10 Min | <br>121.0 ± 15.6<br>119.8 ± 13.2<br>121.6 ± 15.7 | <br>122.4 ±15.8<br>131.3 ± 21.0<br>128.0 ± 20.4 | <br>123.1 ± 7.7<br>126.4 ± 9.8<br>122.9 ± 7.8 | <br>0.60[b]<br>0.24<br>0.072 | — |
| HUT DBP (bpm)[2]:<br>Baseline<br>3 Min<br>10 Min | <br>75.9 ± 15.0<br>78.9 ± 8.5<br>79.9 ± 9.8 | <br>64.7 ± 8.7<br>73.7 ± 12.0<br>73.4 ± 10.3 | <br>67.6 ± 4.8<br>75.7 ± 4.2<br>74.1 ± 5.4 | <br>0.056[b]<br>0.45<br>0.23 | — |
| HUT HR (mmHg)[1]:<br>Baseline<br>3 Min<br>10 Min | <br>76.5 ± 12.9<br>96.9 ± 10.3<br>100.0 ± 9.1 | <br>70.8 ± 9.3<br>83.5 ± 12.9<br>84.7 ± 9.2 | <br>67.6 ± 10.8<br>84.9 ± 13.3<br>84.2 ± 12.8 | <br>0.18<br>0.038*<br>0.0027* | <br><br>IH vs. HC<br>IH vs. NC/HC |
| HUT ΔSBP 3 minutes (mmHg)[1] | -1.2 ± 12.3 | 8.9 ± 10.5 | 3.4 ± 7.7 | 0.10 | |
| HUT ΔDBP 3 minutes (mmHg)[2] | 3.0 ± 9.4 | 9.0 ± 6.2 | 8.1 ± 3.8 | 0.34[b] | — |
| HUT ΔHR[1] | 34.6 ± 18.5 | 26.3 ± 3.4 | 30.8 ± 9.3 | 0.28[b] | — |
| Abnormal HUT: n (%)[2] | 6 (60.0%) | 10 (100%) | 0 (0%) | — | — |
| Valsava baseline to P2 minimum (mmHg)[3] | -6.3 ± 25.1 | — | — | — | — |
| Valsava P2 recovery (mmHg)[3] | 8.1 ± 10.9 | — | — | — | — |
| Valsava P4 overshoot (mmHg)[3] | 18.8 ± 15.6 | 41.4 ± 20.2 | 40.3 ± 18.4 | 0.062[b] | — |
| **Cardiovagal HR variables** | | | | | |
| HRVdb (bpm)[1] | 21.3 ± 5.8 | 24.1 ± 7.8 | 26.8 ± 8.3 | 0.29[b] | — |
| Valsalva Ratio[1] | 2.0 ± 0.34 | 1.7 ± 0.36 | 1.9 ± 0.23 | 0.098 | |
| Abnormal HRVdb or Valsalva Maneuver: n (%) | 0 (0%) | 0 (0%) | 0 (0%) | — | — |
| **Sympathetic cholinergic dysfunction (sudomotor)[4]** | | | | | |
| QSART volume forearm | 0.57 ± 0.87 | — | — | — | — |
| QSART volume proximal leg | 0.73 ± 0.95 | — | — | — | — |
| QSART volume distal leg | 0.62 ± 0.69 | — | — | — | — |
| QSART volume foot | 0.45 ± 0.90 | — | — | — | — |

| | | | | | |
|---|---|---|---|---|---|
| Abnormal QSART: n (%) | 6 (66.7%) | — | — | — | — |
| Length dependent: n (%) | 1 (14.3%) | — | — | — | — |

**Supplementary Table 5. Polysomnography (PSG)-derived sleep macroarchitecture parameters among patients with idiopathic hypersomnia (IH), narcolepsy, and healthy controls (HC) with full PSG data.** Parametric tests were done using a one-way analysis of variance (ANOVA) followed by Turkey HSD or Games-Howell[a] test based on equal variance adjusted for multiple comparisons. Non-parametric tests were done using a Kruskal-Wallis's[b] test followed by Dunn's post hoc test with Bonferroni adjustment for multiple comparisons[b]. Statistical significance (*) is shown for P-value < 0.05. [1] data available only for 3 controls and only an independent t-test was conducted for IH vs. NC. [2] data available for only 10 persons with IH. *Abbreviations*: TST – total sleep time; WASO – wake after sleep onset.

| Variables | IH (n = 11) Mean ± SD Median (IQR) | Narcolepsy (n = 10) Mean ± SD Median (IQR) | Controls (n = 14) Mean ± SD Median (IQR) | P-value | Post-hoc |
|---|---|---|---|---|---|
| **Mean Sleep Onset Latency Testing** | | | | | |
| Mean sleep-onset latency (minutes)[1] | 5.6 ± 2.3<br>4.6 (4.0) | 4.0 ± 1.2<br>3.9 (2.2) | 18.0 ± 2.0<br>18.0 (2.0) | 0.049*[1] | — |
| Number of sleep-onset REM periods[1] | 0.18 ± 0.40<br>0 (0) | 4.0 ± 1.0<br>4.0 (1.7) | 0 | <0.0001* | — |
| **Overnight Polysomnography** | | | | | |
| Sleep-onset latency (minutes)[2] | 20.7 ± 16.9<br>17.0 (10.4) | 10.4 ± 18.2<br>0 (10.0) | 27.1 ± 39.2<br>12.0 (31.6) | 0.11[b] | — |
| REM-sleep latency during (min)[2] | 146.5 ± 107.6<br>100.7 (79.9) | 59.9 ± 57.9<br>72.5 (80.7) | 64.6 ± 30.4<br>66.2 (19.0) | 0.035*[b] | IH vs. HC |
| Sleep efficiency (%)[2] | 89.9 ± 4.2<br>90.0 (5.0) | 76.4 ± 10.6<br>75.0 (13.5) | 81.4 ± 14.2<br>85.2 (13.4) | 0.021*[b] | IH vs. NC |
| TST (min)[2] | 429.0 ± 45.5<br>424.0 (58.0) | 415.4 ± 73.8<br>404.0 (87.5) | 386.1 ± 72.1<br>389.5 (62.2) | 0.28 | — |
| WASO (min)[2] | 27.1 ± 15.1<br>22.5 (27.7) | 117.1 ± 53.2<br>129.0 (89.2) | 66.4 ± 40.7<br>58.5 (20.1) | <0.0001*[a] | IH vs. NC/HC |
| Stage N1 sleep (% TST)[2] | 4.8 ± 2.2<br>4.8 (2.2) | 17.9 ± 7.2<br>16.5 (5.0) | 5.4 ± 3.5<br>4.0 (2.5) | <0.0001*[b] | IH vs. NC<br>NC vs. HC |
| Stage N2 sleep (% TST)[2] | 59.8 ± 14.2<br>62.0 (15.7) | 38.1 ± 9.2<br>39.0 (12.5) | 44.2 ± 7.5<br>45.4 (10.8) | 0.00010* | IH vs. NC/HC |
| Stage N3 sleep (% TST)[2] | 17.0 ± 11.6<br>14.3 (15.3) | 20.3 ± 7.7<br>20.0 (11.0) | 24.0 ± 5.7<br>25.2 (4.3) | 0.16 | — |
| REM sleep (% TST)[2] | 16.9 ± 10.7<br>15.5 (13.1) | 23.9 ± 5.6<br>25.5 (9.5) | 26.3 ± 7.5<br>26.0 (11.1) | 0.028* | IH vs. HC |

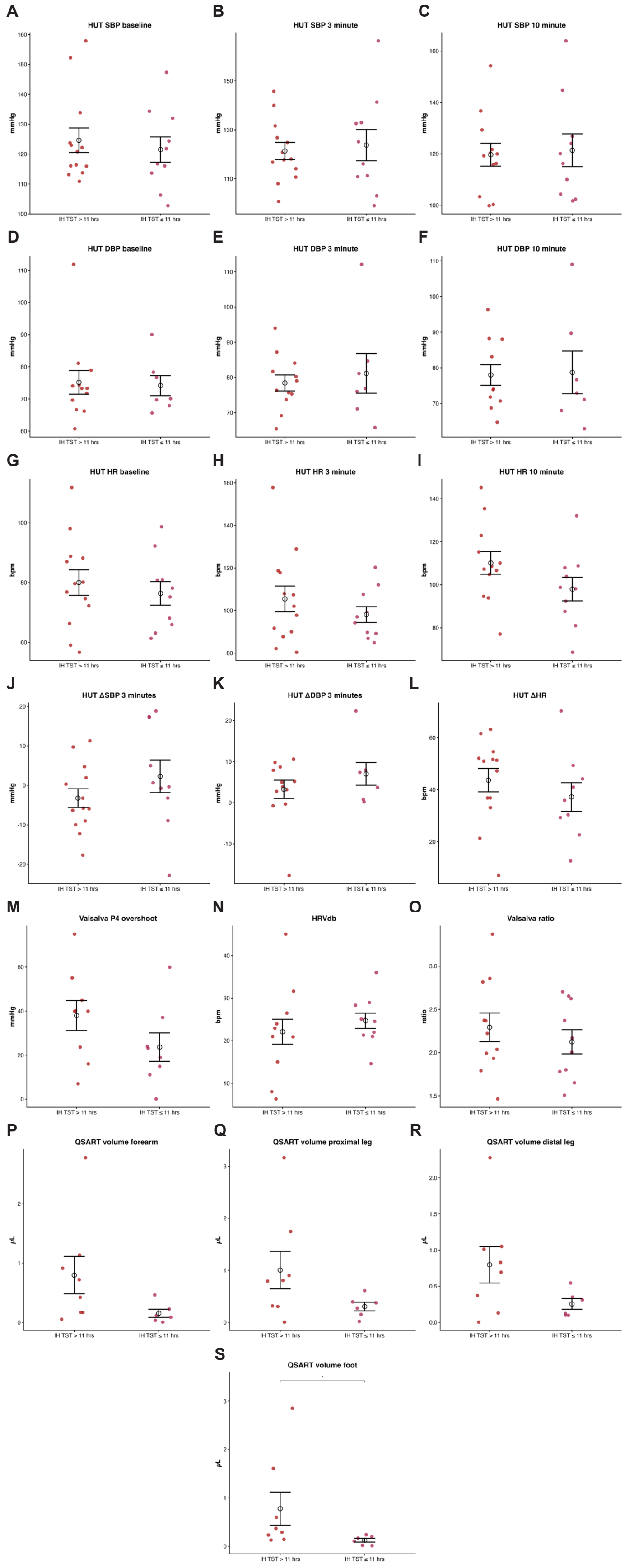

A
HUT SBP baseline
mmHg
B
HUT SBP 3 minute
mmHg
C
HUT SBP 10 minute
mmHg
D
HUT DBP baseline
mmHg
E
HUT DBP 3 minute
mmHg
F
HUT DBP 10 minute
mmHg
G
HUT HR baseline
bpm
H
HUT HR 3 minute
bpm
I
HUT HR 10 minute
bpm
J
HUT ΔSBP 3 minutes
mmHg
K
HUT ΔDBP 3 minutes
mmHg
L
HUT ΔHR
bpm
M
Valsalva P4 overshoot
mmHg
N
HRVdb
bpm
O
Valsalva ratio
ratio
P
QSART volume forearm
μL
Q
QSART volume proximal leg
μL
R
QSART volume distal leg
μL
S
QSART volume foot
μL
IH TST > 11 hrs
IH TST ≤ 11 hrs

**Figure S1. Autonomic reflex testing results for persons with idiopathic hypersomnia stratified by total sleep time (TST) > 11 or ≤ 11** Panels A–C: Systolic blood pressure (SBP) at baseline, 3 minutes, and 10 minutes of head-up tilt (HUT). Panels D–F: Diastolic blood pressure (DBP) at baseline, 3 minutes, and 10 minutes. Panels G–I: Heart rate (HR) at baseline, 3 minutes, and 10 minutes of HUT. Panels J–K: ΔSBP and ΔDBP at 3 minutes of tilt. Panel L: Maximal ΔHR during HUT. Panel M: Valsalva P4 overshoot. Panel N: HRV during deep breathing (HRVdb). Panel O: Valsalva ratio. Each dot represents an individual participant; black bars indicate mean ± SEM.

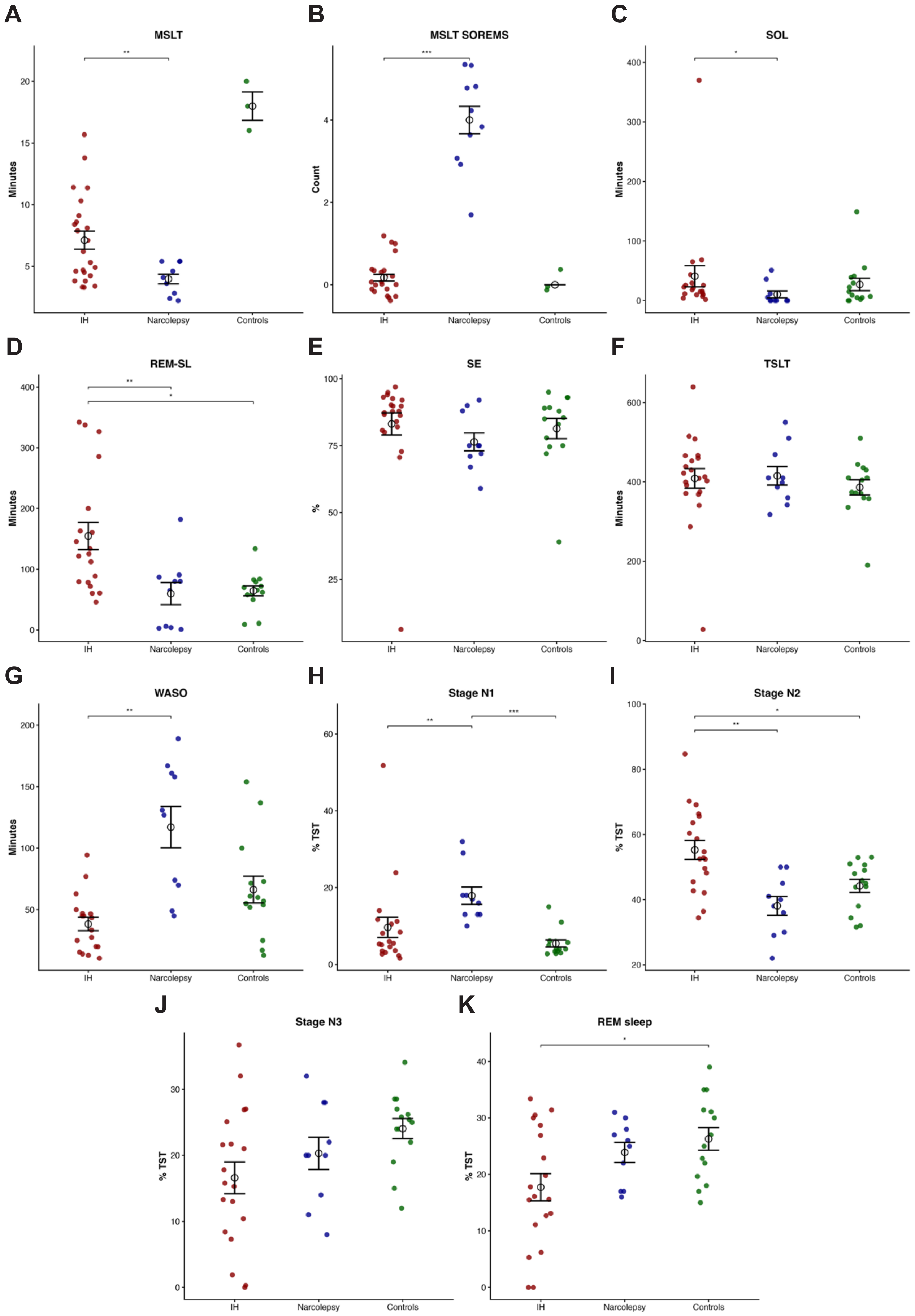
A
MSLT
Minutes
IH
Narcolepsy
Controls
B
MSLT SOREMS
Count
C
SOL
Minutes
D
REM-SL
Minutes
E
SE
%
F
TSLT
Minutes
G
WASO
Minutes
H
Stage N1
% TST
I
Stage N2
% TST
J
Stage N3
% TST
K
REM sleep
% TST

**Figure S2. Sleep and multiple sleep latency test parameters across idiopathic hypersomnia, narcolepsy, and controls.** Panels A–C: mean sleep latency on the multiple sleep latency test (MSLT), number of sleep onset rapid eye movement periods (SOREMs), and sleep onset latency (SOL). Panels D–F: rapid eye movement sleep latency (REM-SL), sleep efficiency (SE), and total sleep time (TST). Panels G–I: wake after sleep onset (WASO), stage N1 (percentage of total sleep time [TST]), and stage N2 (percentage of total sleep time [TST]). Panels J–K: stage N3 (percentage of total sleep time [TST]) and rapid eye movement sleep (percentage of total sleep time [TST]). Each dot represents an individual participant; black bars indicate mean ± SEM.

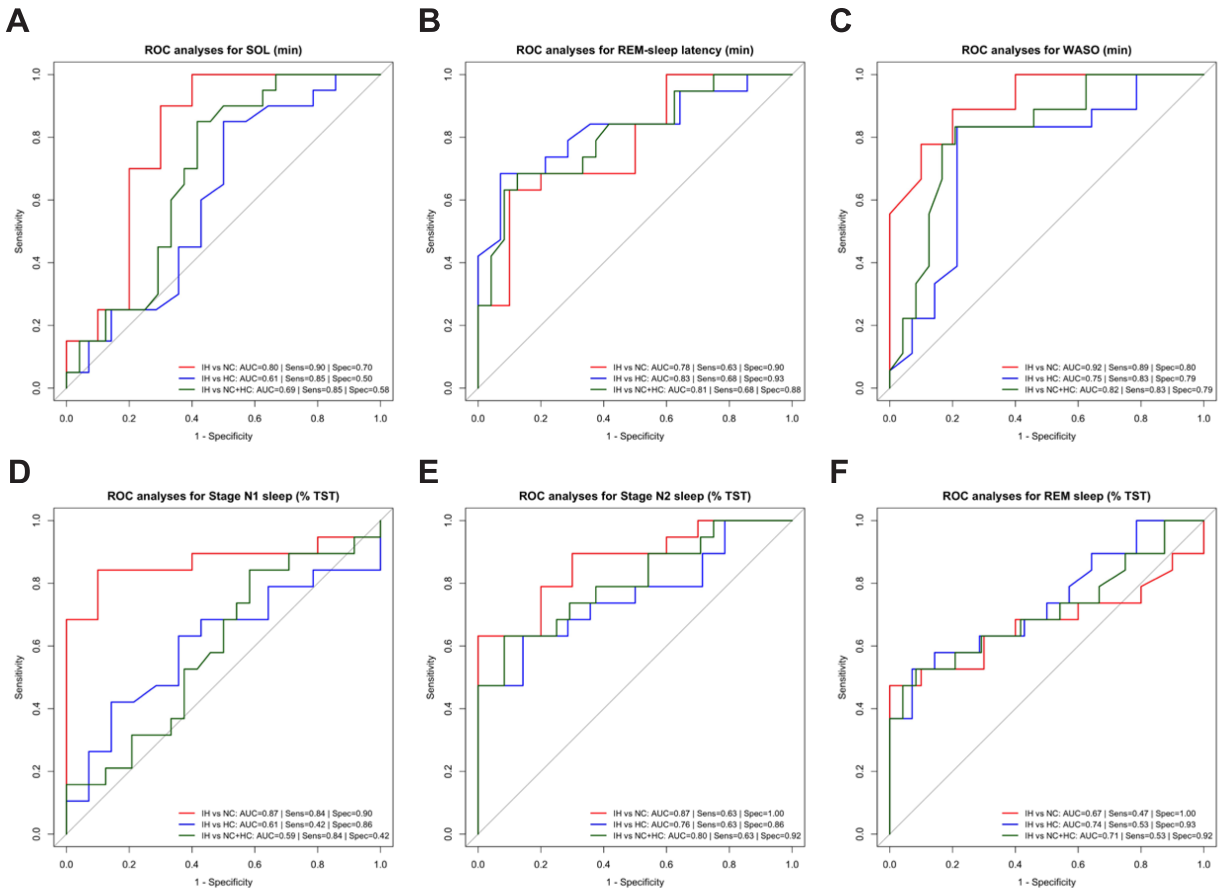


**Figure S3. Receiver operating characteristic analyses for sleep parameters across idiopathic hypersomnia, narcolepsy, and controls**. Panels A–C: sleep onset latency (SOL), rapid eye movement sleep latency (REM-SL), and wake after sleep onset (WASO). Panels D–F: stage N1 sleep (percentage of total sleep time [TST]), stage N2 sleep (percentage of total sleep time [TST]), and rapid eye movement sleep (percentage of total sleep time [TST]). Curves represent comparisons between idiopathic hypersomnia vs narcolepsy, idiopathic hypersomnia vs controls, and idiopathic hypersomnia vs combined narcolepsy and controls, with corresponding area under the curve (AUC), sensitivity, and specificity values shown.

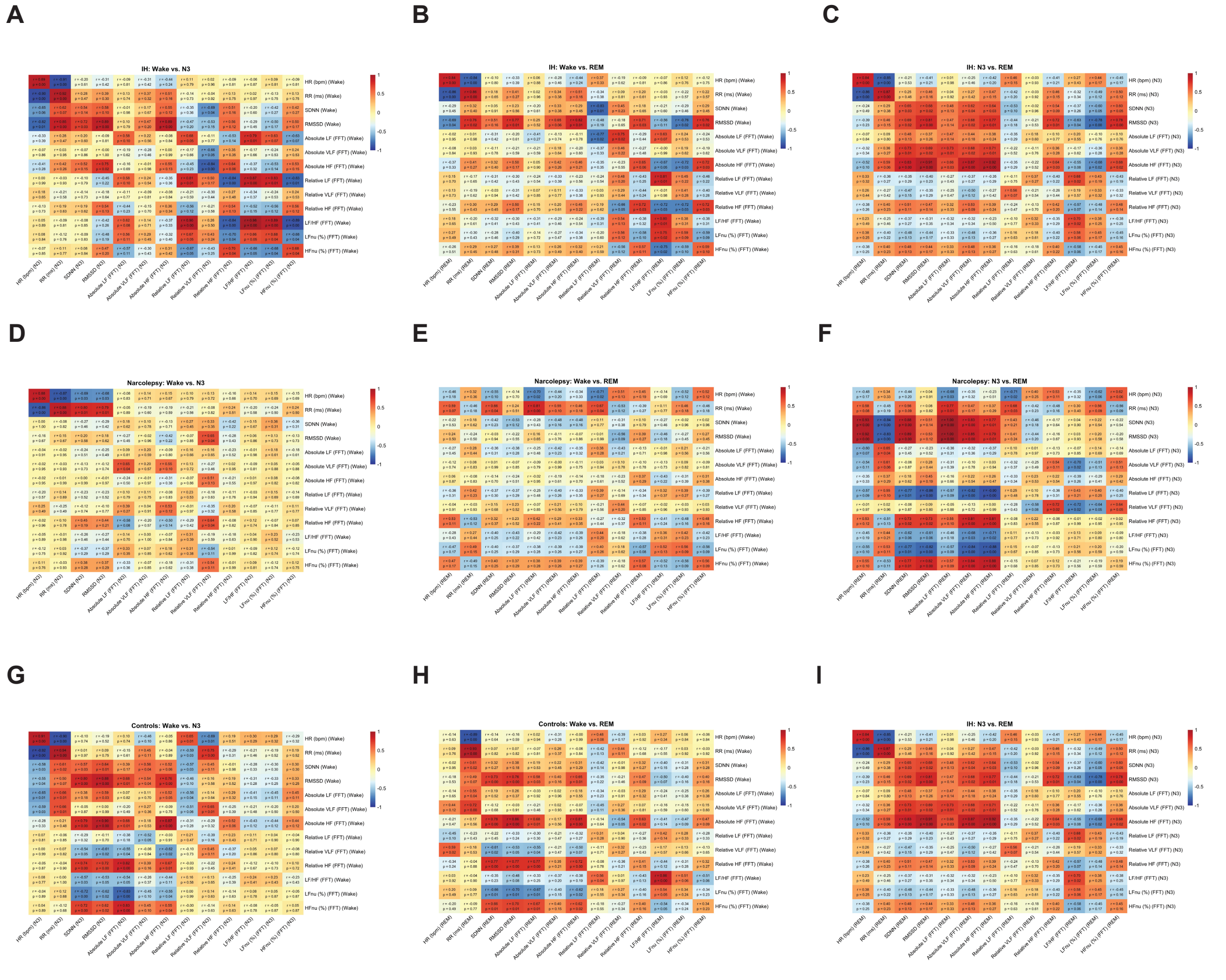


**Figure S3. Correlation heatmaps of heart rate and heart rate variability parameters across sleep stages.** Panels A–C: idiopathic hypersomnia (IH), showing wake vs. N3 (A), wake vs. REM (B), and N3 vs. REM (C). Panels D–F: narcolepsy type 1 (NT1), showing wake vs. N3 (D), wake vs. REM (E), and N3 vs. REM (F). Panels G–I: healthy controls, showing wake vs. N3 (G), wake vs. REM (H), and N3 vs. REM (I). Heatmaps display pairwise correlations between heart rate and time- and frequency-domain HRV measures, including SDNN, RMSSD, and spectral indices (LF, HF, LF/HF), with color scales indicating correlation coefficients (red = positive, blue = negative).
